# XPro1595, a Selective Soluble TNF Neutralizer, in Early Alzheimer’s Disease with Inflammation (ADi): Results from the Phase 2 MINDFuL Trial

**DOI:** 10.1101/2025.09.24.25336496

**Authors:** Judith Jaeger, Kim A. Staats, Sarah Barnum, Parris Pope, Lisle Kingery, Melanie Buitendyk, Sharon Cohen, Malú Gámez Tansey, Raymond J Tesi, CJ Barnum

## Abstract

**Background:** Alzheimer’s disease (AD) pathophysiology involves chronic neuroinflammation driven by tumor necrosis factor (TNF). XPro1595 selectively neutralizes pathological soluble TNF while preserving neuroprotective transmembrane TNF signaling, offering a mechanistically distinct approach from current amyloid-targeting therapies.

**Methods:** The MINDFuL trial was a Phase 2, multicenter, randomized, double-blind, placebo-controlled study conducted across 35 centers in eight countries. Participants aged 50-85 years with Early AD (mild cognitive impairment or mild dementia) and elevated inflammatory biomarkers received weekly subcutaneous injections of XPro1595 (1.0 mg/kg) or placebo for 24 weeks. The primary endpoint was the Early Mild Alzheimer’s Cognitive Composite (EMACC). A prospectively defined biomarker-enriched population included amyloid-positive participants with a high inflammatory burden (**ADi**), defined by the presence of ≥2 inflammatory markers.

**Results:** Of 206 randomized participants, 200 comprised the modified intent-to-treat (mITT) population and 100 comprised the ADi population. In the mITT analysis, no significant difference was observed in the EMACC (p=0.672). However, the ADi population showed promising signals across multiple clinical endpoints favoring XPro1595, including the EMACC (Cohen’s d = 0.27), Neuropsychiatric Inventory (NPI, d = 0.23), particularly the subfactor agitation/hyperactivity (d = 0.37), and a patient-reported outcome: Goal Attainment Scale (GAS, d = 0.18). These results were consistent with biomarker changes that favored XPro1595, as plasma pTau217 (d = -0.18) and GFAP (d = -0.19) were attenuated in the ADi population. In addition, many effect sizes were larger when assessing participants with higher drug exposure. Notably, no amyloid-related imaging abnormalities (ARIA) were observed, even though ∼70% of the participants were APOE ε4 carriers, and one-third had baseline microbleeds.

**Conclusions:** While XPro1595 did not meet the primary endpoint in the overall population, participants with biologically-defined AD and high inflammatory burden demonstrated consistent trends toward cognitive benefit and neuropsychiatric improvement. The complete absence of ARIA distinguishes XPro1595 from amyloid-targeting therapies and supports its potential for use in combination strategies and potentially for those at a higher risk of ARIA. These findings support selective TNF inhibition as a promising therapeutic approach for persons with AD and elevated inflammatory burden.

## Introduction

Alzheimer’s disease (AD) and related dementias represent one of the most urgent and rapidly escalating global health challenges of the 21st century, affecting more than 44 million individuals worldwide and projected to surpass $1 trillion USD in annual cost by 2050. Therapeutic efforts over the past several decades have primarily focused on amyloid-β, a defining pathological hallmark of AD. Despite the success of recently approved anti-amyloid therapies, their modest effects on disease progression and risk of adverse events, particularly amyloid-related imaging abnormalities (ARIA), underscore the urgent need for new mechanistic approaches.

Innate immune dysfunction and chronic inflammation are increasingly recognized as central drivers of AD pathophysiology. Tumor necrosis factor (TNF), a master regulator of inflammatory signaling, plays a critical role in processes ranging from amyloid accumulation^1^ to synaptic loss^2^ and neuronal death^3^. TNF exists in two bioactive forms: soluble TNF (sTNF), which promotes neuroinflammation and cell death, and transmembrane TNF (tmTNF)^4^, which facilitates anti-inflammatory and neuroprotective glial functions. Conventional anti-TNF therapies indiscriminately block both forms, often leading to compromised immune defense and demyelination^5–7^. XPro1595 was developed to address this limitation by selectively neutralizing sTNF while preserving TNF-mediated protective signaling. Through this mechanism, XPro1595 reduces pathological inflammation without impairing essential immune and glial functions^8^. Preclinical studies have demonstrated that selective sTNF blockade with XPro1595 decreases glial activation, enhances debris clearance, and restores synaptic plasticity^2^. A Phase 1b study in AD extended these findings, showing dose-dependent reductions in cerebrospinal fluid biomarkers of neuroinflammation and neurodegeneration (manuscript in preparation).

The present Phase 2 proof-of-concept study, MINDFuL, was designed to test the hypothesis that modulating inflammation with XPro1595 has a measurable impact on cognitive function in Early AD. Recognizing that immune-targeted interventions may require trial designs distinct from amyloid-focused approaches, this study incorporated two defining features. First, biomarker enrichment ensured the inclusion of participants with evidence of inflammation, thereby aligning patient biology with the mechanism of action of XPro1595. Second, the primary outcome was the Early Mild Alzheimer’s Cognitive Composite (EMACC), an empirically derived measure composed of validated clinical neuropsychological tests specifically optimized to detect cognitive changes that occur in early disease; therefore, it is well suited for Phase 2 signal detection. These design features were integrated with established clinical and biomarker measures to ensure comparability with traditional AD trials while also offering critical insights into how to evaluate immune-targeted therapies in AD more effectively.

## Methods

### Study Design and Participants

MINDFuL (NCT05318976, EudraCT 2023-505396-71-00) was a Phase 2, multicenter, randomized, double-blind, placebo-controlled trial of XPro1595 in participants with Early AD and signs of inflammation. “Early AD” was defined as mild cognitive impairment (MCI) or mild Alzheimer’s dementia, consistent with NIA-AA Stage 3–4 disease^9^. The trial was conducted across 35 research centers in eight countries (Australia, Canada, Czech Republic, France, Germany, Poland, Spain, and the United Kingdom) between February 28, 2022, and April 2024. The study protocol was approved by the ethics committee of each site, and all participants or their legal representatives provided written informed consent.

Participants were 50–85 years old with a diagnosis of MCI or mild AD dementia (consistent with the McKhann criteria^10^ and a Clinical Dementia Rating global score of 0.5–1.0) at screening. Key inclusion criteria evolved slightly due to protocol amendments (Supplemental Table 2), but the core target population comprised individuals with confirmed amyloid pathology and biomarkers of inflammation. Originally, eligible participants were required to have amyloid positivity (as determined by historical documentation or blood analysis) and ≥2 inflammatory biomarkers from a predefined panel (described below). During recruitment, operational constraints led to protocol amendments permitting the inclusion of participants without amyloid confirmation (when timely amyloid testing was unavailable) and with at least one elevated inflammatory biomarker. Once the operational restraints were resolved, confirmation of amyloid positivity became mandatory for inclusion again (Supplemental Table 2). All participants had a Mini-Mental State Examination (MMSE) score ≥ 22 at screening, and, when applicable, an Everyday Cognition (ECog) memory subscale item mean > 1.5, consistent with mild cognitive impairment or very mild dementia severity. The key exclusion criteria included evidence of other neurological disorders and clinically significant lesion(s), either of which could confound or indicate a dementia diagnosis other than AD, and the chronic use of immunosuppressive therapies that could obscure the potential effect of XPro1595 (Supplemental Table 1 for the full list of inclusion and exclusion criteria.)

### Inflammation Biomarker Enrichment

A key inclusion criterion was the presence of at least one of the following biomarkers of inflammation:

- High sensitivity C-reactive Protein (CRP) > 1.5 mg/L
- Erythrocyte Sedimentation Rate (ESR) > 10 mm/h
- Hemoglobin A1C (HbA1C) > 6DCCT%, or
- At least 1 Apolipoprotein E4 (APOE4) allele

Inflammatory biomarkers were used to enrich the trial for two reasons: First, selecting participants with inflammation-related biomarkers aligns their pathophysiology with the drug mechanism of action. Second, participants with biomarkers of inflammation are more likely to experience faster disease progression^11^, which could allow for the observation of a treatment response within a shorter period of time. The inflammatory biomarkers were chosen based on meeting at least one of the following criteria: (1) evidence linking them to cognitive decline or association with AD^11^ (2) their ability to predict treatment response to anti-inflammatory or anti-TNF therapies^12^, and (3) their routine use by primary care physicians and ease-of-measurement in local clinical laboratories.

### Randomization and Masking

A total of 721 individuals were screened, of whom 206 met the eligibility criteria and were randomized in a 2:1 ratio to receive XPro1595 (1.0 mg/kg) or placebo, respectively. Randomization was stratified by clinical stage (MCI vs. mild AD) and sex to ensure balance between the treatment arms. Study treatment (XPro1595 or placebo) was administered in a blinded manner. Participants, caregivers, investigators, and study staff were blinded to the treatment assignment. XPro1595 and placebo were provided as identical-appearing solutions for once-weekly subcutaneous injections (administered by a study nurse or caregiver).

### Study Design

The treatment period was 24 weeks of double-blind therapy, followed by a 4-week safety follow-up after the final dose. Biological measures, including blood and imaging, were obtained at baseline, 12, and 24 weeks. The EMACC and a delayed verbal memory task (the ISRL) were administered every 6 weeks for 24 weeks. All other clinical rating measures were completed every 12 weeks (baseline, 12 weeks, and 24 weeks; Supplementary Figure 1). Participants were seen at the clinic when assessments were performed. Visits where participants only received drug administration could be performed at the clinic or at the participant’s home. Adherence was monitored using injection logs and drug accountability counts. An independent Data Monitoring Committee reviewed unblinded safety data at periodic intervals to ensure the well-being of the participants.

### Outcome Measures

Cognitive performance was assessed using the ***Early/Mild Alzheimer’s Cognitive Composite (EMACC),*** which served as the primary efficacy endpoint. The EMACC is a composite of six widely used and well-validated neuropsychological tests that assess memory, verbal fluency, and executive function. It was empirically developed from four different aging cohorts and was specifically designed to be optimally sensitive to change in Early AD. The tests in the EMACC include the International Shopping List Test (ISLT) (verbal learning), digit symbol coding (information processing speed), digits forward and digits backward subtests of the Wechsler Adult Intelligence Scale (working memory), the Letter Fluency and Category Fluency tests from the Delis-Kaplan Executive Function System (verbal fluency), and Trail Making Test (TMT) parts A and B (executive function). The EMACC has substantial overlap with the Preclinical Alzheimer’s Cognitive Composite (PACC), a composite of neuropsychological tests widely used in secondary prevention trials, before the onset of objective cognitive impairment. The EMACC score is a z-transformed composite that was standardized using the baseline performance of all study participants per population. Baseline distributions were previously reported^13^ with individual and composite measures normally distributed in the overall study sample. The EMACC was administered at screening (for training), baseline, and every 6 weeks (weeks 6, 12, 18, and 24). A higher EMACC score indicates better cognitive performance.

The key secondary endpoint was the ***Clinical Dementia Rating Sum of Boxes (CDR-SB***), based upon a clinician-administered semi-structured interview of the participant and informant^14^. The CDR-SB (range, 0–18) reflects the degree of impairment in cognitive and functional skills across six domains (3 cognitive domains: memory, orientation, judgment and problem solving; and 3 functional domains: community affairs, home and hobbies, and personal care), with higher scores indicating greater impairment. In addition to CDR-SB, the cognitive and functional domains were evaluated separately. The CDR rater was blinded to the EMACC, NPI, and ADCS-ADL scales. The CDR-SB and subsequent clinical scales were administered at baseline, week 12, and week 24.

The informant version of the ***Measurement of Everyday Cognition (ECog***) scale^15^ was included to measure observed changes in everyday functioning associated with cognitive impairment. The ECog is a 39-item questionnaire in which the study partner rates the participant’s change in his/her ability to complete cognitively relevant everyday activities when compared to 10 years prior (i.e. prior to illness onset). The activities rated reflect changes in memory, language, visual spatial/perceptional, planning, organization, and divided attention skills. Higher scores reflect a worsening ability to complete tasks and, therefore, a greater decline since prior to illness onset.

The ***Neuropsychiatric Inventory (NPI)***^16^ measures neuropsychiatric and behavioral symptoms of dementia via a caregiver interview. The NPI assesses the frequency and severity of 12 behavioral symptoms (delusions, hallucinations, agitation/aggression, depression, anxiety, euphoria, apathy, disinhibition, irritability, aberrant motor behavior, nighttime disturbances, and appetite changes), with higher scores indicating worse neuropsychiatric problems. Subfactor analyses were conducted on agitation/hyperactivity (agitation/aggression, elation/euphoria, disinhibition, and irritability/lability items) and on mood/depression (depression/dysphoria, anxiety, apathy/indifference, sleep, and appetite and eating disorders items). These subfactors were determined by literature review and defined *a priori*.

Functional ability was measured using the study partner report on the ***Alzheimer’s Disease Cooperative Study Activities of Daily Living scale***. Depending on the protocol version and patient stage, either the ADCS-MCI-ADL (for MCI)^17^ or the ADCS-ADL (for mild AD)^18^ was used. The ADCS-ADL assesses both basic ADLs (fundamental self-care tasks such as dressing and bathing) and instrumental ADLs (more complex tasks of daily living such as managing finances and preparing meals). The ADCS-MCI-ADL focuses solely on instrumental ADL skills, as basic activities of daily living are not typically impacted at the MCI stage of disease.

The ***Goal Attainment Scale (GAS)*** is an individualized outcome measure that quantifies the effects of an intervention based on personalized goals^19,20^. With a trained interviewer, participants and their study partners set treatment goals at baseline that were meaningful to them, and relevant to their life circumstances and potential response to the treatment. Initially developed to evaluate mental health services, GAS offers a semi-structured approach to individualized, patient-centric outcome measurement. It has been used in a variety of indications (e.g., Alzheimer disease, cerebral palsy, autism) and is a highly responsive method of capturing participants lived experience with their condition and treatment. The achievement of each goal is rated on a 5-point attainment scale (-2, -1, 0, +1, +2) to allow standardized scoring of personalized outcomes. A participant’s overall goal attainment is quantified using a formula that considers the number of goals that have been set and the extent to which they are correlated with each other. At each site, the same rater performed the GAS assessment for a specific participant throughout the study.

The ***International Shopping List Test, Delayed Recall (ISRL)***^21^ is one component of an auditory verbal learning test in which a respondent is first given three trials to recall a 12-item shopping list (ISLT immediate recall) and then, after an approximate 20-minute delay, is asked to recall all of the words they can remember from the list of 12 words (ISLT delayed recall). The ISRL raw score is the number of words that the participant can freely recall after the delay. This free recall ISRL portion of the ISLT is especially sensitive to detecting memory impairments in AD^21^.

Two blood-based disease-relevant biomarkers were measured to explore XPro1595’s impact on AD-related pathology and neuroinflammation. Blood samples for pTau217 and GFAP were collected at baseline and at Week 24 (end-of-treatment), processed to plasma, and analyzed by Quanterix Corp using the Simoa ALZpath p-Tau 217 and Simoa GFAP Advantage PLUS kits.

### Safety Assessments

Safety and tolerability were evaluated via adverse event (AE) monitoring, vital signs, physical and neurological exams, laboratory tests (hematology, chemistry, liver enzymes, etc.), and electrocardiograms. Brain MRI was performed at screening and repeated at Week 24 to monitor for ARIA (edema or microhemorrhages) and other structural changes. An independent neuroradiologist adjudicated MRI for ARIA and other abnormalities. All AEs and serious adverse events (SAEs) were recorded and coded by system organ class, and their severity and relationship to study drug was assessed by investigators blind to treatment. An independent Data Monitoring Committee (DMC) assessed ongoing safety at prespecified time points. Participant safety was continuously monitored by an external medical monitoring team blinded to study treatment, who reviewed all AEs and consulted with PIs as needed. All participants who received any amount of XPro1595 or Placebo are included in the Safety Analysis Set (SAF). All safety data were derived from the SAF.

### Analysis Populations

The Safety Analysis Set (SAF) included all participants who received at least one dose of study medication (XPro1595 or placebo). The modified Intent-to-Treat (mITT) population included all randomized participants who received at least one dose and had at least one post-baseline efficacy assessment. The mITT population (n = 200) was the primary population for efficacy analyses and reflects the broad enrolled sample, including participants under the expanded inclusion criteria. The prospectively defined ADi population, was analyzed separately. The ADi population (n = 100) consisted of those mITT participants who met the original criteria of amyloid positivity plus ≥2 inflammatory biomarkers at baseline. This population represents the target population for which the trial was powered. Efficacy analyses were performed in both the mITT and ADi groups. We also assessed the ‘dose compliant’ participants, who received at least 21 mg/kg of study drug throughout the study (XPro1595 or placebo), which were n=79 of the ADi. The trial’s planned sample size (n=201) was estimated via power calculations anticipating a moderate effect on EMACC in the originally recruited population (similar to ADi). As 50% of the mITT met the original enrichment criteria, statistical power for the primary endpoint was reduced.

### Statistical Analysis

Efficacy was analyzed using a mixed-effects model for repeated measures (MMRM). The MMRM included change-from-baseline scores at each post-baseline visit as the dependent variable, with fixed effects for treatment group, visit (categorical), and treatment-by-visit interaction. The model also included the following covariates: indication (MCI/mAD), sex (M/F), confirmed amyloid status (yes/no), age, number of inflammatory biomarkers (1, 2, 3, 4), country, and the baseline value of the endpoint being tested. An unstructured covariance matrix was used to model within-participant covariance errors. From the MMRM, least-squares mean (LSM) changes from baseline for each group and the LSM difference between XPro1595 and placebo at Week 24 were estimated, along with 90% confidence intervals (CI) and p-values for the treatment difference. An alpha level (two-sided α=0.10, corresponding to a one-sided 5% test) was prespecified for hypothesis testing at Week 24, recognizing the Phase 2 exploratory nature. The primary comparison was the EMACC change at Week 24 (mITT). Key secondary endpoints were tested in a hierarchical manner to control family-wise error rate (EMACC followed by CDR-SB), but given the primary outcome was not significant, mITT secondary results are reported with nominal p-values. To evaluate the ADi population, we used effect size as the primary metric. Effect size, measured by Cohen’s *d*, is well-suited for small samples and allows comparisons across different measures (e.g., cognitive tests and biomarkers). Unlike p-values, which indicate the likelihood of results being due to chance, effect size reflects clinical relevance and is commonly used for signal detection in Phase 2 studies. A coherent pattern of directional findings with a promising signal would be supportive of a biologically and/or clinically meaningful effect that may have been missed as a result of inadequate study power. We defined a promising signal as a minimum effect size of 0.2, where XPro1595 outperformed placebo on multiple endpoints aligned with our hypothesis and the drug’s mechanism of action. Signal detection was based on the effect size difference in LS mean change from baseline (MMRM model) between XPro1595 and placebo at 24 weeks, ensuring results were meaningful, relevant, and appropriate for the trial’s design and objectives. Safety data were summarized in the SAF population with descriptive statistics. All analyses were performed using SAS v9.4.

### Data Availability

The datasets generated and/or analyzed during the current study are not publicly available due to confidentiality and intellectual property restrictions associated with the developmental stage of XPro1595. Limited, de-identified data that support the findings of this study may be made available to qualified researchers, subject to review and a data sharing agreement with INmune Bio, Inc.

## Results

### Participant Disposition and Baseline Characteristics

The double-blind treatment phase ran from February 2022 to April 2024. Of 721 individuals screened, 207 were eligible and randomized (140 allocated to XPro1595 and 67 to placebo; Fig. 1). The Safety (SAF) population included 206 participants who received ≥1 dose (one randomized placebo participant did not start treatment). The mITT population comprised 200 participants (134 XPro1595, 66 placebo) with at least one post-baseline efficacy measurement. The ADi population included 100 participants from mITT that were amyloid-positive and had ≥2 inflammatory biomarkers at baseline (72 XPro1595, 28 placebo).

**Figure 1.**
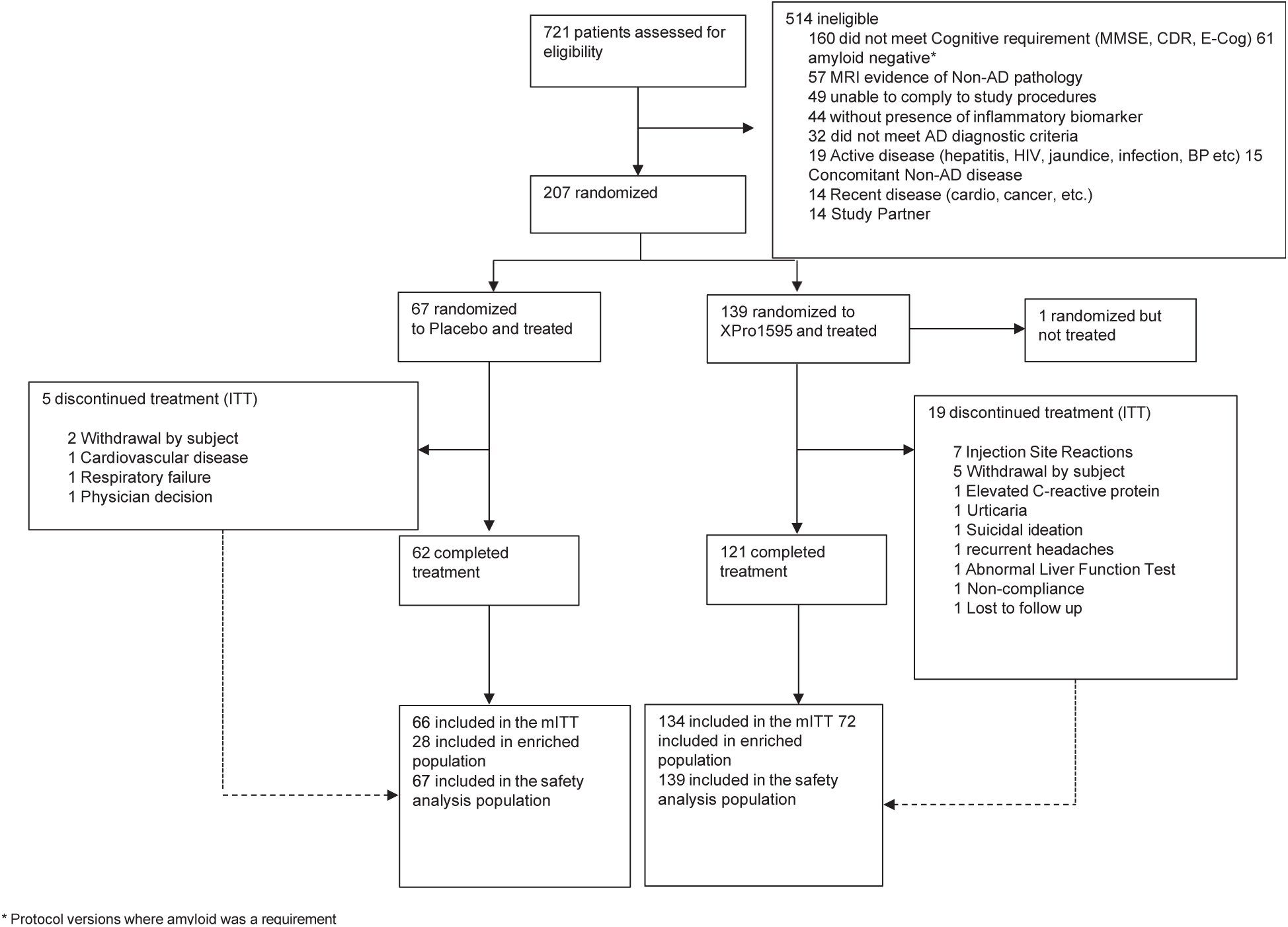
Participant Flow and Disposition in MINDFuL. CONSORT diagram showing screening, randomization, treatment allocation, and completion rates. Of 721 individuals screened, 207 were randomized 2:1 to XPro1595 (n=140) or placebo (n=67). The modified intent-to-treat (mITT) population included 200 participants (134 XPro1595, 66 placebo), while the biomarker-enriched ADi population comprised 100 participants (72 XPro1595, 28 placebo) with confirmed amyloid positivity and ≥2 inflammatory biomarkers. Overall completion rate was 88.4%, with slightly higher discontinuation in the XPro1595 arm (13.7% vs 7.5%).

Overall, 24 participants (11.6%) did not complete the 24-week treatment period. Discontinuations were slightly higher in the XPro1595 arm: 19/139 (13.7%) vs 5/67 (7.5%) in placebo. The most common reason for study withdrawal on XPro1595 was adverse events (12 participants, see safety), with smaller numbers due to personal choice (5), non-compliance (1), or loss to follow-up (1). In the placebo group, 3 discontinued due to adverse events and 2 withdrew consent/elected to stop.

Baseline demographic and clinical characteristics were comparable between treatment groups (Table 1). The mean participant age was 72.4 +/-6.5 years, with 51% female. At baseline, approximately 45% of participants were classified as MCI, and 55% as mild AD. Participants in the XPro1595-treated arm had more advanced disease at baseline in both mITT and ADi populations, as indicated by a greater percentage of mAD (vs MCI), longer time since diagnosis, higher CDR-SB scores, and poorer performance on cognitive tasks (EMACC and ISRL). The XPro1595-treated group had a higher average number of elevated inflammatory biomarkers at baseline compared to the placebo group, in both mITT and ADi. Although these imbalances were modest, they suggest that the XPro1595-treated participants, on average, began the study with a greater disease burden and higher levels of inflammation than the placebo-treated participants.

**Table 1.**
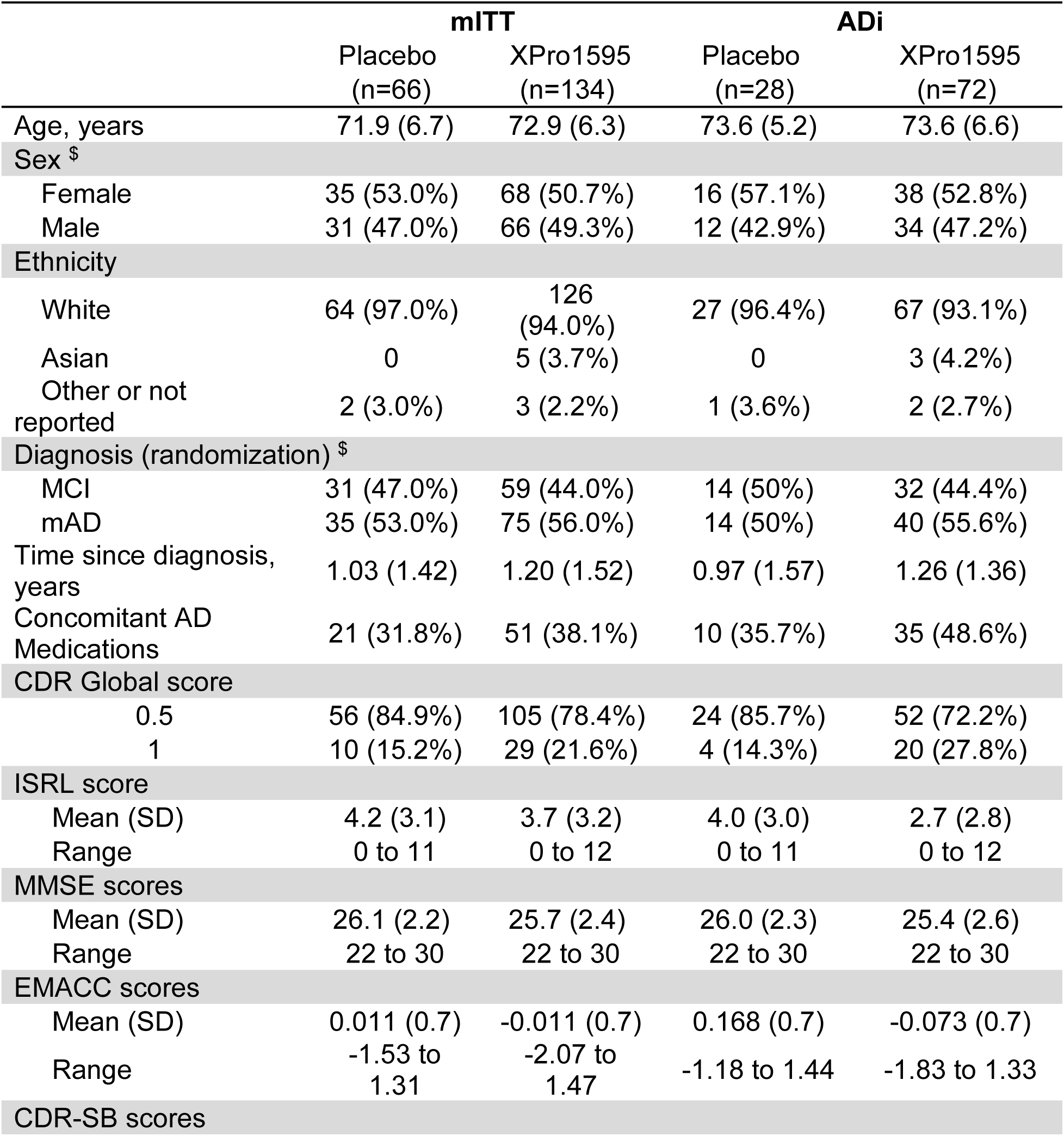
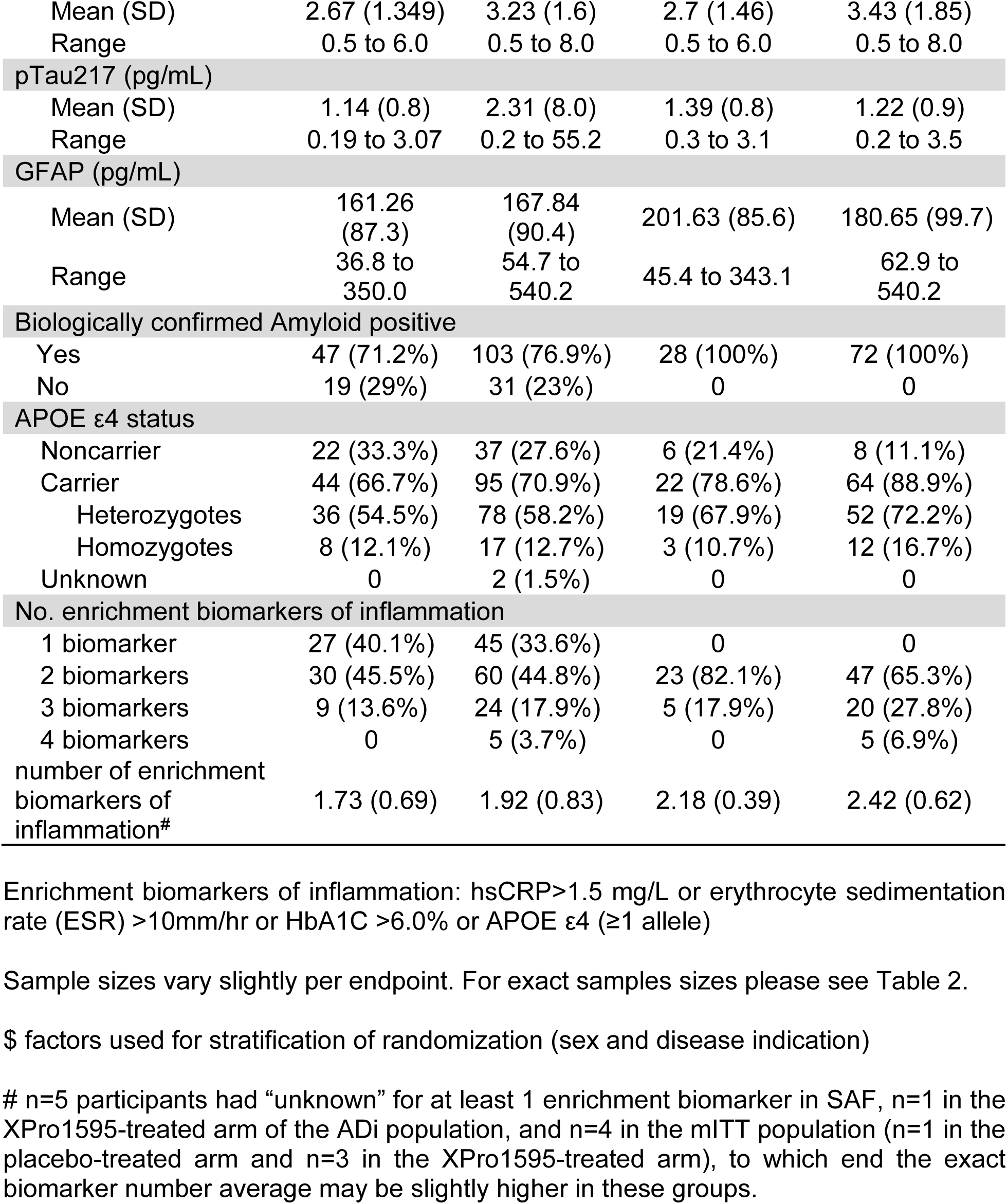
Baseline Demographics and Clinical Characteristics. Demographic and clinical characteristics at baseline for mITT and ADi populations. The XPro1595 group had slightly more advanced disease burden at baseline, including higher proportions of mild AD (vs MCI), longer time since diagnosis, and greater inflammatory biomarker burden. Despite randomization stratification, modest imbalances favored the placebo group, particularly in the ADi population. Values shown as mean (SD) for continuous variables and n (%) for categorical variables.

|  | mITT |  | ADi |  |
| --- | --- | --- | --- | --- |
|  | Placebo<br>(n=66) | XPro1595<br>(n=134) | Placebo<br>(n=28) | XPro1595<br>(n=72) |
| Age, years | 71.9 (6.7) | 72.9 (6.3) | 73.6 (5.2) | 73.6 (6.6) |
| Sex <sup>§</sup> |  |  |  |  |
| Female | 35 (53.0%) | 68 (50.7%) | 16 (57.1%) | 38 (52.8%) |
| Male | 31 (47.0%) | 66 (49.3%) | 12 (42.9%) | 34 (47.2%) |
| Ethnicity |  |  |  |  |
| White | 64 (97.0%) | 126 (94.0%) | 27 (96.4%) | 67 (93.1%) |
| Asian | 0 | 5 (3.7%) | 0 | 3 (4.2%) |
| Other or not reported | 2 (3.0%) | 3 (2.2%) | 1 (3.6%) | 2 (2.7%) |
| Diagnosis (randomization) <sup>§</sup> |  |  |  |  |
| MCI | 31 (47.0%) | 59 (44.0%) | 14 (50%) | 32 (44.4%) |
| mAD | 35 (53.0%) | 75 (56.0%) | 14 (50%) | 40 (55.6%) |
| Time since diagnosis, years | 1.03 (1.42) | 1.20 (1.52) | 0.97 (1.57) | 1.26 (1.36) |
| Concomitant AD Medications | 21 (31.8%) | 51 (38.1%) | 10 (35.7%) | 35 (48.6%) |
| CDR Global score |  |  |  |  |
| 0.5 | 56 (84.9%) | 105 (78.4%) | 24 (85.7%) | 52 (72.2%) |
| 1 | 10 (15.2%) | 29 (21.6%) | 4 (14.3%) | 20 (27.8%) |
| ISRL score |  |  |  |  |
| Mean (SD) | 4.2 (3.1) | 3.7 (3.2) | 4.0 (3.0) | 2.7 (2.8) |
| Range | 0 to 11 | 0 to 12 | 0 to 11 | 0 to 12 |
| MMSE scores |  |  |  |  |
| Mean (SD) | 26.1 (2.2) | 25.7 (2.4) | 26.0 (2.3) | 25.4 (2.6) |
| Range | 22 to 30 | 22 to 30 | 22 to 30 | 22 to 30 |
| EMACC scores |  |  |  |  |
| Mean (SD) | 0.011 (0.7) | -0.011 (0.7) | 0.168 (0.7) | -0.073 (0.7) |
| Range | -1.53 to 1.31 | -2.07 to 1.47 | -1.18 to 1.44 | -1.83 to 1.33 |
| CDR-SB scores |  |  |  |  |
| Mean (SD) | 2.67 (1.349) | 3.23 (1.6) | 2.7 (1.46) | 3.43 (1.85) |
| Range | 0.5 to 6.0 | 0.5 to 8.0 | 0.5 to 6.0 | 0.5 to 8.0 |
| pTau217 (pg/mL) |  |  |  |  |
| Mean (SD) | 1.14 (0.8) | 2.31 (8.0) | 1.39 (0.8) | 1.22 (0.9) |
| Range | 0.19 to 3.07 | 0.2 to 55.2 | 0.3 to 3.1 | 0.2 to 3.5 |
| GFAP (pg/mL) |  |  |  |  |
| Mean (SD) | 161.26 (87.3) | 167.84 (90.4) | 201.63 (85.6) | 180.65 (99.7) |
| Range | 36.8 to 350.0 | 54.7 to 540.2 | 45.4 to 343.1 | 62.9 to 540.2 |
| Biologically confirmed Amyloid positive |  |  |  |  |
| Yes | 47 (71.2%) | 103 (76.9%) | 28 (100%) | 72 (100%) |
| No | 19 (29%) | 31 (23%) | 0 | 0 |
| APOE ε4 status |  |  |  |  |
| Noncarrier | 22 (33.3%) | 37 (27.6%) | 6 (21.4%) | 8 (11.1%) |
| Carrier | 44 (66.7%) | 95 (70.9%) | 22 (78.6%) | 64 (88.9%) |
| Heterozygotes | 36 (54.5%) | 78 (58.2%) | 19 (67.9%) | 52 (72.2%) |
| Homozygotes | 8 (12.1%) | 17 (12.7%) | 3 (10.7%) | 12 (16.7%) |
| Unknown | 0 | 2 (1.5%) | 0 | 0 |
| No. enrichment biomarkers of inflammation |  |  |  |  |
| 1 biomarker | 27 (40.1%) | 45 (33.6%) | 0 | 0 |
| 2 biomarkers | 30 (45.5%) | 60 (44.8%) | 23 (82.1%) | 47 (65.3%) |
| 3 biomarkers | 9 (13.6%) | 24 (17.9%) | 5 (17.9%) | 20 (27.8%) |
| 4 biomarkers | 0 | 5 (3.7%) | 0 | 5 (6.9%) |
| number of enrichment biomarkers of inflammation<sup>#</sup> | 1.73 (0.69) | 1.92 (0.83) | 2.18 (0.39) | 2.42 (0.62) |

Enrichment biomarkers of inflammation: hsCRP>1.5 mg/L or erythrocyte sedimentation rate (ESR) >10mm/hr or HbA1C >6.0% or APOE ε4 (≥1 allele)
Sample sizes vary slightly per endpoint. For exact samples sizes please see Table 2.
\$ factors used for stratification of randomization (sex and disease indication)

### Primary Cognitive Outcome (EMACC)

In the mITT population, the primary analysis showed no statistically significant difference in EMACC performance change between XPro1595 and placebo at 24 weeks (Table 2). Mean change from baseline was minimal in both arms, with no observable decline in the placebo group. The MMRM-estimated least-squares mean (LSM) difference in EMACC (XPro1595 – placebo) was –0.018 (SE 0.041; 90% CI: –0.086 to +0.051; p = 0.672). The absence of treatment effect appears to result from the unexpected lack of decline in the placebo group (Supplementary Fig. 2). In contrast, the placebo-treated participants in the ADi population exhibited the anticipated decline (Supplementary Fig. 2). Among the ADi population, EMACC change from baseline scores in XPro1595 treated participants compared to placebo had an LSM difference of +0.086 (90% CI: –0.015 to +0.186; p = 0.1594), corresponding to Cohen’s d = 0.27. This suggests a small-to-moderate treatment benefit, with visible separation between treatment groups at Week 24 (Fig. 2). Post hoc exploration of EMACC subtests within the ADi population indicated the treatment effect was not driven by a single measure.

**Figure 2.**
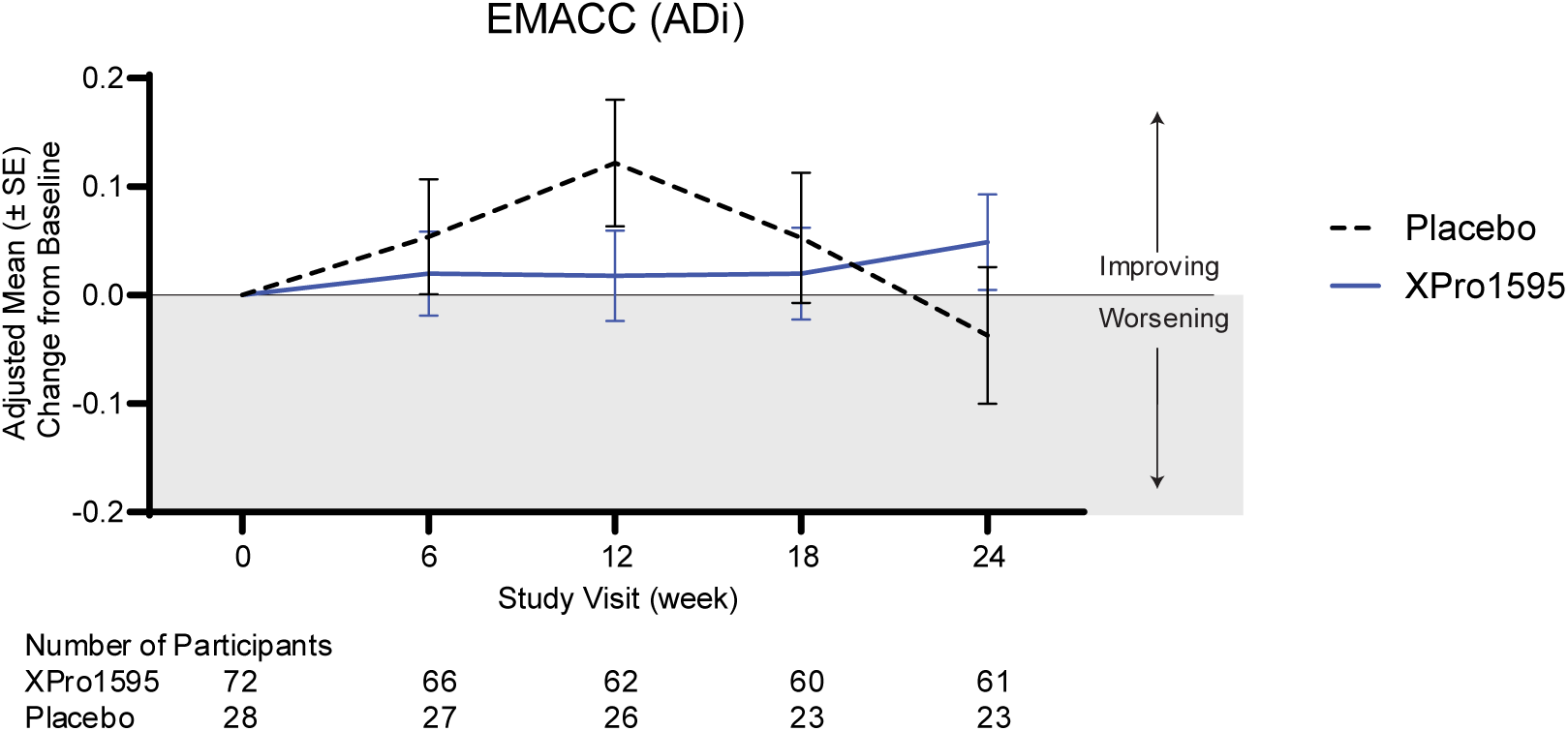
Primary Cognitive Outcome (EMACC) in the ADi population. Change from baseline in Early Mild Alzheimer’s Cognitive Composite (EMACC) scores over 24 weeks. ADi population demonstrating separation between treatment groups by Week 24 when the placebo arm begins to decline (Cohen’s d=0.27). Higher EMACC scores indicate better cognitive performance. Data presented as Adjusted least-squares mean changes +/-SEM.

**Table 2.** Summary of Study Population results. Summarizing table of the results on MINDFuL at week 24. Number of participants per population (in n below) are the number of participants included in the analysis at 24 weeks (earlier time points may have slightly varying number of participants included).

| Study population results at week 24 |  | N |  |  |  | 90% CI |  |  |  |  |
| --- | --- | --- | --- | --- | --- | --- | --- | --- | --- | --- |
| Endpoint | Analysis set | Placebo | XPro1595 | LSM | SE | lower | upper | p-value | cohen's d | Interpretation (based on cohen's d) |
| EMACC | mITT | 58 | 111 | -0.02 | 0.04 | -0.09 | 0.05 | 0.67 | -0.04 | Favorable trend in participants with high inflammatory burden |
|  | ADi | 23 | 61 | 0.09 | 0.06 | -0.01 | 0.19 | 0.16 | **0.27** |  |
|  | Dose compliant ADi | 22 | 51 | 0.07 | 0.07 | -0.04 | 0.19 | 0.28 | **0.21** |  |
| ISRL | mITT | 57 | 110 | 0.05 | 0.33 | -0.49 | 0.59 | 0.88 | 0.00 | Directional trend in participants with high inflammatory burden |
|  | ADi | 23 | 61 | 0.41 | 0.42 | -0.29 | 1.11 | 0.33 | 0.16 |  |
|  | Dose compliant ADi | 22 | 51 | 0.11 | 0.44 | -0.62 | 0.84 | 0.80 | 0.05 |  |
| CDRSB | mITT | 59 | 120 | -0.11 | 0.18 | -0.42 | 0.19 | 0.55 | -0.06 | Directional trend in participants that had greater drug exposure |
|  | ADi | 24 | 67 | -0.08 | 0.31 | -0.59 | 0.43 | 0.79 | -0.05 |  |
|  | Dose compliant ADi | 22 | 56 | -0.22 | 0.32 | -0.76 | 0.31 | 0.49 | -0.13 |  |
| CDRSB (Cognition) | mITT | 59 | 120 | -0.09 | 0.11 | -0.28 | 0.10 | 0.44 | -0.08 | Directional trend in participants that had greater drug exposure |
|  | ADi | 24 | 67 | -0.06 | 0.19 | -0.37 | 0.25 | 0.74 | -0.06 |  |
|  | Dose compliant ADi | 22 | 56 | -0.13 | 0.20 | -0.47 | 0.21 | 0.52 | -0.12 |  |
| CDRSB (Function) | mITT | 59 | 120 | 0.04 | 0.11 | -0.14 | 0.22 | 0.72 | 0.04 | No trend |
|  | ADi | 24 | 67 | 0.02 | 0.18 | -0.27 | 0.32 | 0.90 | 0.02 |  |
|  | Dose compliant ADi | 22 | 56 | -0.03 | 0.18 | -0.33 | 0.27 | 0.85 | -0.04 |  |
| NPI | mITT | 59 | 120 | -0.90 | 0.78 | -2.18 | 0.39 | 0.25 | -0.11 | Favorable trend in participants with high inflammatory burden and greater drug exposure |
|  | ADi | 24 | 67 | -1.62 | 1.25 | -3.71 | 0.47 | 0.20 | **-0.23** |  |
|  | Dose compliant ADi | 22 | 56 | -1.90 | 1.39 | -4.22 | 0.42 | 0.18 | **-0.27** |  |
| NPI (Agitation/Hyperactivity) | mITT | 59 | 120 | -0.40 | 0.25 | -0.82 | 0.02 | 0.11 | -0.17 | Favorable trend in participants with high inflammatory burden |
|  | ADi | 24 | 67 | -0.90 | 0.42 | -1.55 | -0.17 | 0.04 | **-0.37** |  |
|  | Dose compliant ADi | 22 | 56 | -0.90 | 0.48 | -1.66 | -0.06 | 0.08 | **-0.34** |  |
| NPI (Mood/Depression) | mITT | 59 | 120 | 0.20 | 0.23 | -0.20 | 0.55 | 0.45 | 0.09 | Unfavorable Directional trend in participants with high inflammatory burden |
|  | ADi | 24 | 67 | 0.20 | 0.33 | -0.31 | 0.79 | 0.47 | 0.16 |  |
|  | Dose compliant ADi | 22 | 56 | 0.30 | 0.34 | -0.30 | 0.84 | 0.44 | 0.17 |  |
| Ecog | mITT | 58 | 119 | 0.07 | 0.08 | -0.06 | 0.20 | 0.38 | 0.07 | No trend |
|  | ADi | 24 | 65 | 0.12 | 0.12 | -0.08 | 0.32 | 0.33 | 0.14 |  |
|  | Dose compliant ADi | 22 | 54 | 0.06 | 0.12 | -0.14 | 0.26 | 0.63 | 0.08 |  |
| pTau217 | mITT | 41 | 81 | 0.08 | 0.04 | 0.01 | 0.15 | 0.07 | 0.19 | Favorable trend in participants with high inflammatory burden and greater drug exposure |
|  | ADi | 19 | 49 | -0.04 | 0.06 | -0.13 | 0.05 | 0.44 | -0.18 |  |
|  | Dose compliant ADi | 14 | 43 | -0.07 | 0.06 | -0.18 | 0.03 | 0.26 | **-0.26** |  |
| GFAP | mITT | 38 | 85 | -0.01 | 0.04 | -0.08 | 0.06 | 0.75 | -0.03 | Favorable trend in participants with high inflammatory burden and greater drug exposure |
|  | ADi | 18 | 50 | -0.06 | 0.06 | -0.17 | 0.05 | 0.36 | -0.19 |  |
|  | Dose compliant ADi | 14 | 44 | -0.07 | 0.07 | -0.20 | 0.05 | 0.33 | **-0.22** |  |
| GAS | mITT | 64 | 126 | 1.63 | 1.80 | -1.91 | 5.17 | 0.37 | 0.14 | Favorable trend in participants with high inflammatory burden |
|  | ADi | 28 | 69 | 2.00 | 2.50 | -3.00 | 7.00 | 0.42 | 0.18 |  |
|  | Dose compliant ADi | na | na | na | na | na | na | na | na |  |

(+) cohen's d indicates trend for XPro1595 for EMACC, ISRL, and GAS. (-) cohen's d indicates trend for XPro1595 for CDR-SB, NPI, ECog, pTau217 and GFAP. Where the absolute value of cohen's d <0.1 = No trend; Where the absolute value of cohen's d ≥0.1 = potential trend. Where the absolute value of cohen's d >0.2 = trend. na - not available.

### Secondary Neuropsychiatric and Functional Outcomes

Global measures of cognition and function, including CDR-SB, did not differ between treatment arms over 24 weeks in the mITT population (LSM difference –0.11, SE 0.185; 90% CI –0.417 to 0.195; p = 0.55; d = –0.06) or in the ADi population (LSM difference – 0.08; SE 0.307; 90% CI –0.593 to 0.426; p = 0.79; d = –0.05). In the dose compliant ADi population, the effect size increased modestly (LSM difference –0.22, SE = 0.32, 90% CI –0.76 - 0.31, p=0.45, d = –0.13), suggesting treatment exposure may influence efficacy. In addition, assessing only the cognitive domains of the CDR-SB, indicates that the minimal directional effect of XPro1595 is larger for cognition domains than those of function (Fig. 3)

**Figure 3.**
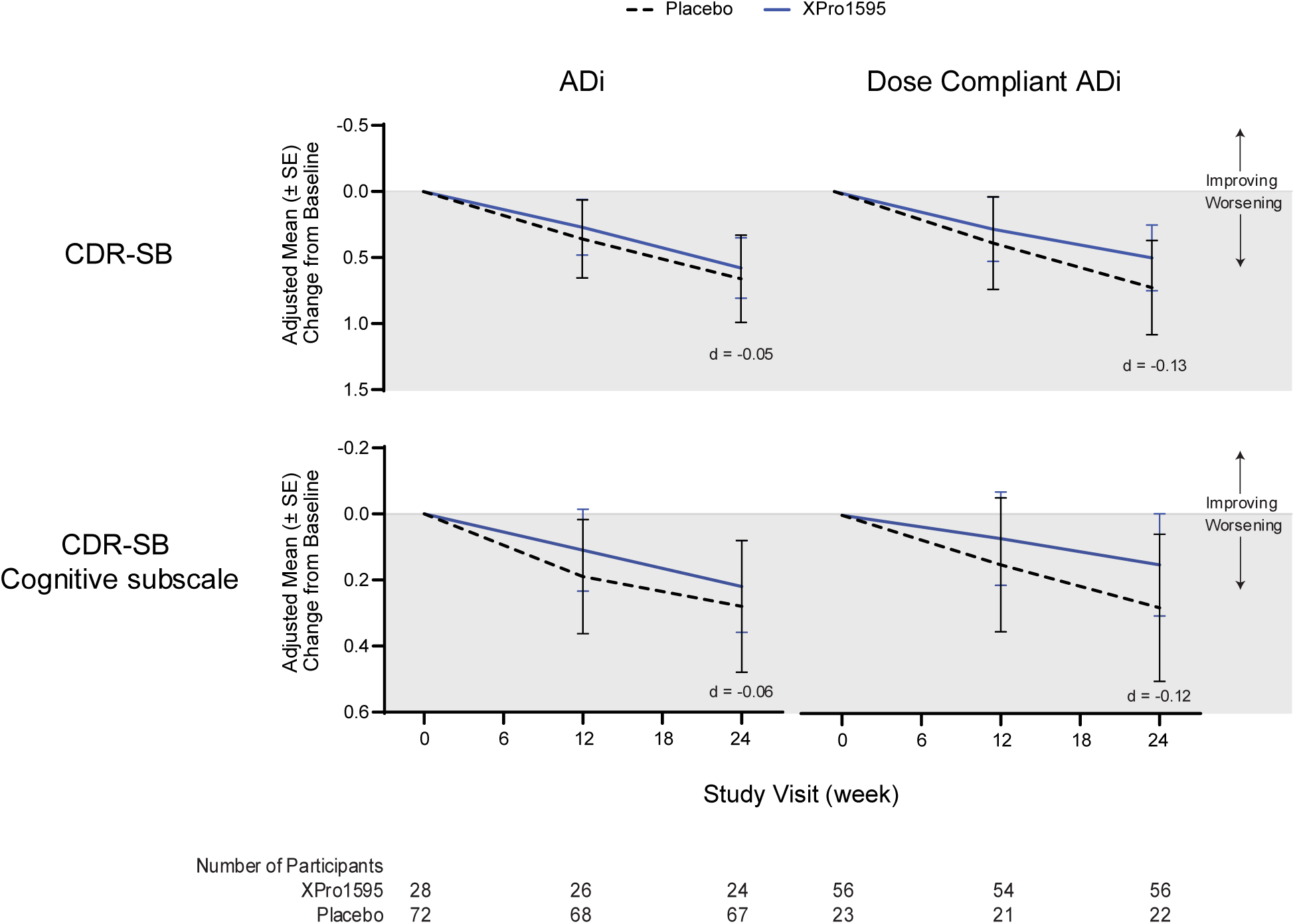
CDR-SB in the ADi and Dose compliant ADi population. Change from baseline in CDR-SB over 24 weeks in the full CDR-SB scale and the subscale of cognition. The dose compliant ADi population demonstrates increased separation between treatment groups by Week 24. Negative CDR-SB scores indicate better cognitive performance. Data presented as Adjusted least-squares mean changes +/-SEM.

Moreover, treatment with XPro1595 was associated with consistent positive trends in neuropsychiatric symptoms across analysis populations (Table 2). In the mITT population, NPI scores indicated symptom improvement in XPro1595 treated participants as compared to worsening in placebo participants (LSM difference –0.9; 90% CI –2.18 to 0.39; d = –0.11; p = 0.250). The effect was more pronounced in the ADi population (LSM difference –1.6; 90% CI –3.71 to 0.47; d = –0.23; p = 0.200), largely reflecting worsening outcomes in placebo-treated participants (Fig. 4). Cumulative dose, and thus drug exposure, appeared important as the positive trend for XPro1595 was greater in participants who received at least 21 mg/kg of study drug (’dose compliant’) with an effect size of –0.17 (LSM difference -1.4, SE =0.837, 90% CI –2.76 to 0.02, p = 0.104). This trend was even greater in the dose compliant ADi population with an effect size of –0.27 (LSM difference –1.9, 90% CI –4.22 to 0.42, p=0.176).

**Figure 4.**
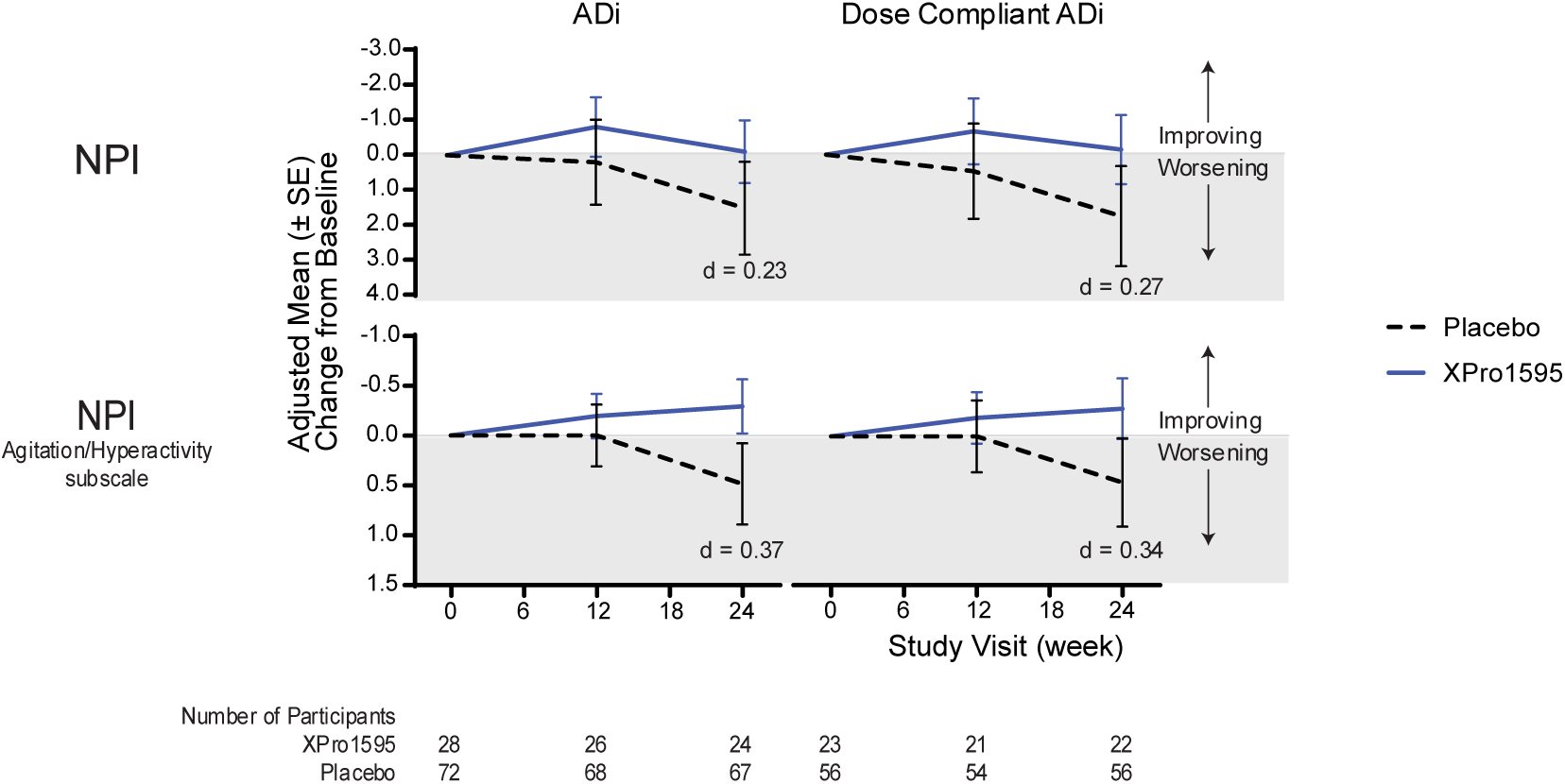
Neuropsychiatric Inventory (NPI) in the ADi and Dose compliant ADi population. A trend for XPro1595 was observed in the NPI in both the ADi (cohen’s d = 0.23) and Dose compliant ADi (cohen’s d = 0.27) populations; with placebo-treated participants experiencing worsening symptoms. Subfactor-specific analysis of agitation/hyperactivity in ADi (cohen’s d=-0.37) and Dose complaint ADi populations (cohen’s d = 0.34), respectively. Negative NPI scores indicate fewer neuropsychiatric symptoms. Data presented as Adjusted least-squares mean changes +/-SEM.

Subfactor-level analyses showed the largest benefit on the agitation/hyperactivity factor (Fig. 4) with an effect size of -0.17 in the mITT population (LSM difference –0.4; 90% CI -0.82 to 0.02, p = 0.114) and -0.37 in the ADi population (LSM difference –0.9; 90% CI – 1.55 to –0.17; p = 0.041). Acknowledging the clinical relevance of agitation for AD patients and caregivers, this finding is notable.

Everyday Cognition (ECog) informant-reports showed no treatment effect (Table 2) in mITT (LSM difference = 0.07; SE = 0.08, 90% CI –0.06 - 0.20, p=0.38, d=0.07) or in ADi (LSM difference 0.06, SE= 0.12, 90% CI –0.14-0.26, p=0.63 d= 0.08), though ceiling effects were noted at baseline. ADCS-ADL outcomes were uninterpretable due to version differences, administration errors, and ceiling effects.

Goal Attainment Scaling (GAS) showed numerically greater improvements with XPro1595 (Table 2) in ADi (LSM difference 2.00, SE=2.50, 90% CI -3.00 to 7.00, p=0.42, d=0.18) than in mITT (LSM difference 1.63, SE = 1.80, 90% CI –1.91 to 5.17, p=0.37, d=0.14), suggesting participants receiving active drug were somewhat more likely to meet personal goals. GAS results may capture quality-of-life effects not reflected in conventional outcome measures.

The delayed recall component of the word list learning task (ISRL) is a well-validated measure of episodic memory, capturing both acquisition and retention of new information. Because impaired delayed recall is a hallmark of Alzheimer’s disease, ISRL is informative for assessing treatment-related effects on memory. In the mITT population, no difference was observed between treatment groups (LSM difference = 0.1, SE = 0.33, 90% CI = - 0.49 to 0.59, p = 0.8779, d = 0). In the ADi population, a positive trend was noted for XPro1595 (LSM difference = 0.4, SE = 0.42, 90% CI = -0.29 to 1.11, p = 0.3296, d = 0.16).

### Biomarker Outcomes

Plasma ***phosphorylated tau at threonine-217 (pTau217)*** was measured as a marker of Alzheimer’s pathology; pTau217 is a highly specific blood biomarker that correlates with amyloid PET positivity and disease progression^22^. The biomarker analyses revealed a divergent pattern depending on baseline inflammatory status. In the mITT population, XPro1595 treatment was associated with a modest rise in plasma pTau217 relative to placebo (LSM difference 0.08; 90% CI 0.007 to 0.155; p = 0.07; d = 0.19). By contrast, in the ADi population, XPro1595 attenuated the typical 24-week increase observed in placebo-treated participants (LSM difference –0.04; 90% CI –0.135 to 0.050; p = 0.443; d = –0.18; Fig. 5), which was even larger when assessing the dose compliant ADi (LSM difference –0.07, SE=0.06, 90% CI -0.018 to 0.03, p=0.26, d= -0.26). Plasma ***glial fibrillary acidic protein (GFAP)*** was measured as a marker of neuroinflammation and astroglial activation; elevated GFAP reflects reactive astrocytosis around Aβ plaques and is associated with cognitive decline in preclinical AD^23^. A similar pattern as with pTau217 was observed in plasma GFAP, as in the mITT, differences were negligible (LSM difference –0.01; 90% CI –0.084 to 0.057; p = 0.749; d = –0.03). In ADi, XPro1595 reduced the expected increase in GFAP relative to placebo (LSM difference –0.06; 90% CI –0.167 to 0.048; p = 0.356; d = -0.19; Fig. 5) and this effect is larger with increased dose exposure (dose compliant ADi: LSM difference –0.07, SE=0.07, 90% CI –0.20 to 0.05, p=0.33, d= -0.22. Together, these biomarker findings support the hypothesis that XPro1595’s anti-inflammatory mechanism may mitigate downstream tau and astrocytic pathology specifically in participants with elevated inflammation.

**Figure 5.**
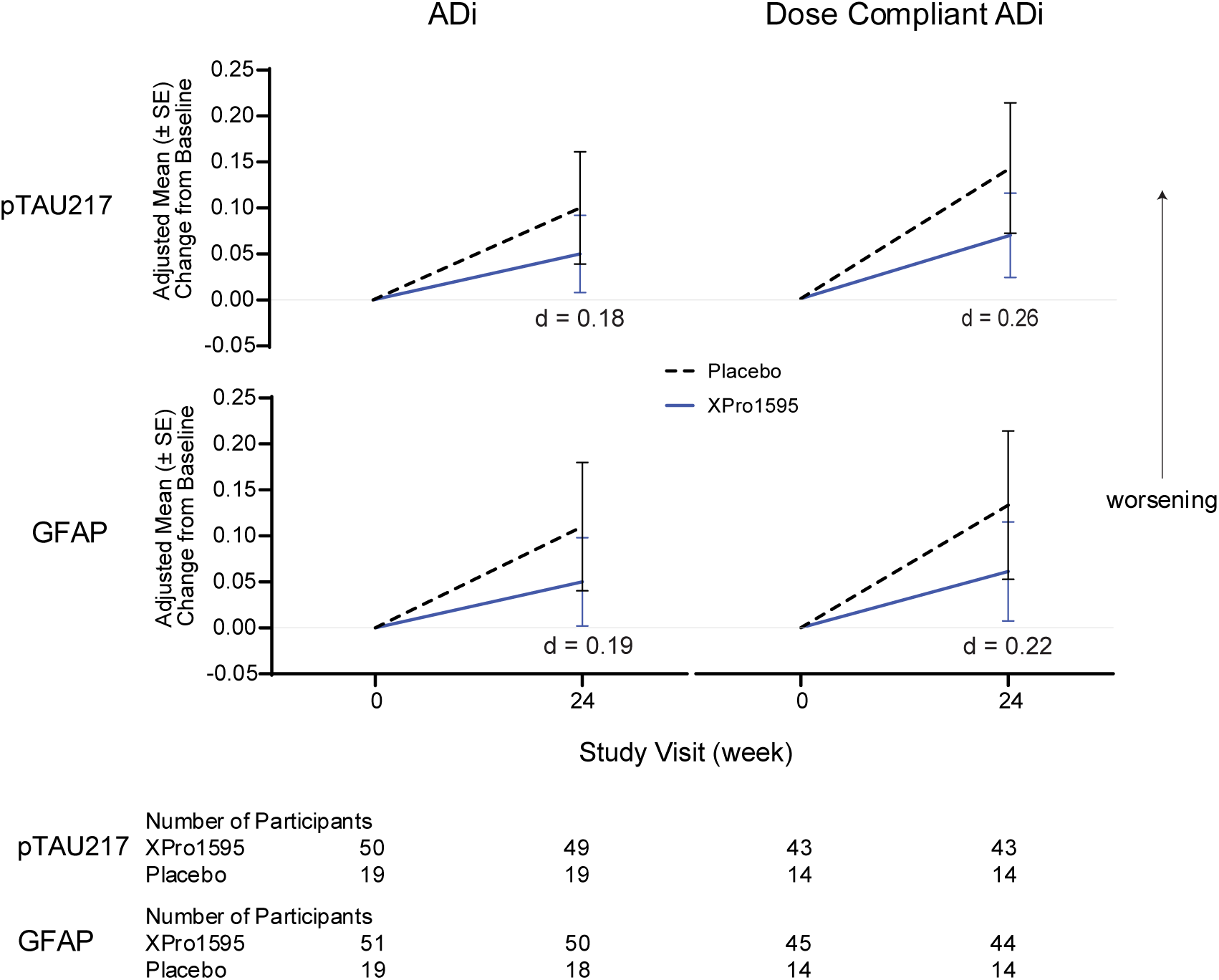
Biomarker Changes from Baseline to Week 24 in the ADi and Dose compliant ADi populations. Treatment with XPro1595 attenuated the increase in plasma pTau217 observed in placebo participants within ADi (cohen’s d = –0.18) and dose-compliant Adi (cohen’s d = –0.26) groups. Similar effects were observed for plasma GFAP, with XPro1595 reducing its trajectory in both the ADi (cohen’s d = –0.19) and dose-compliant ADi groups (cohen’s d = –0.22). Data are shown as Adjusted least-squares mean changes ± SEM.

**Figure 6.**
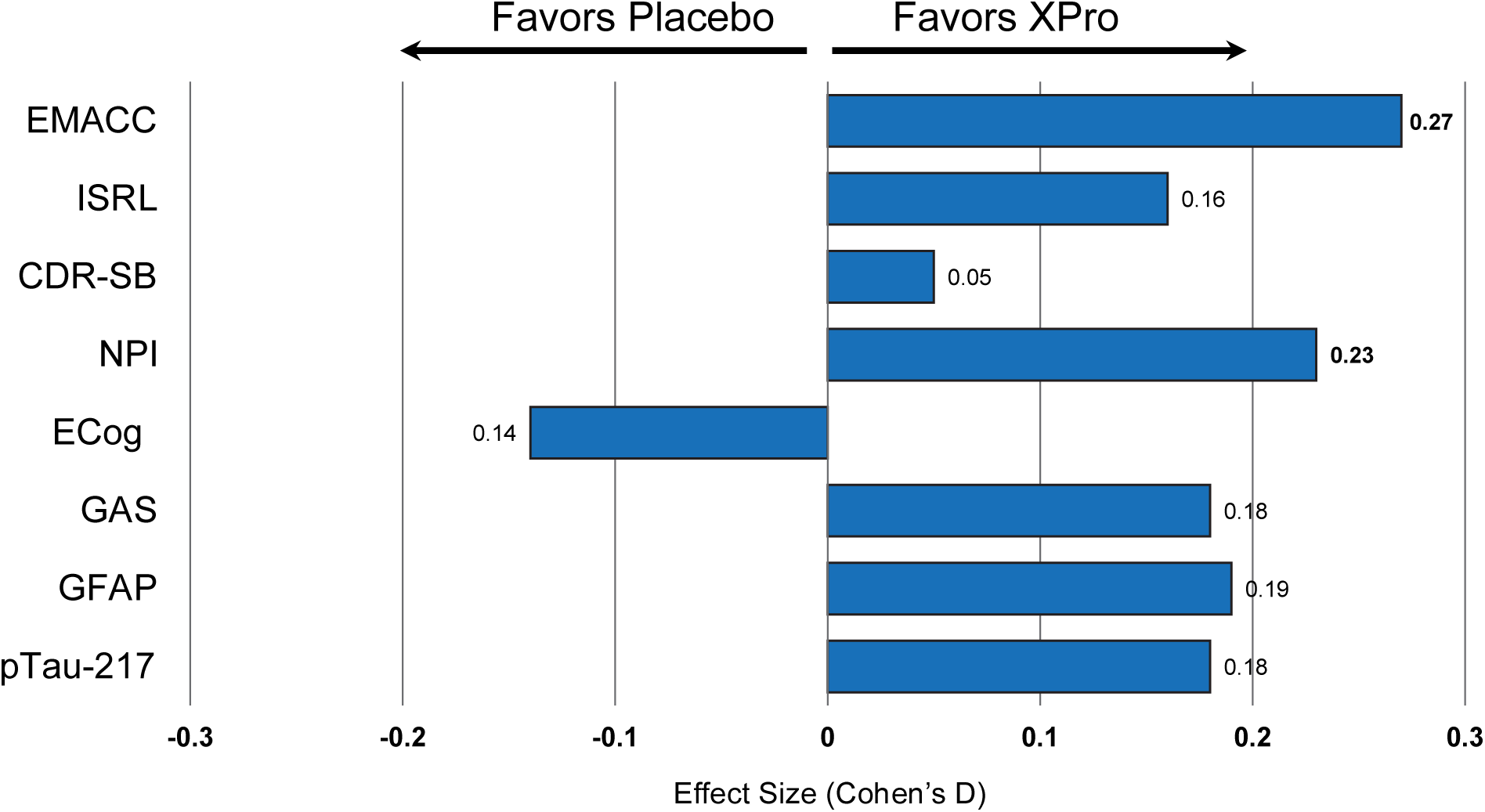
Summary of Treatment Effects Across Endpoints in the ADi Population. Forest plot displaying effect sizes (Cohen’s d) for key efficacy endpoints in the ADi population (n=100). XPro1595 demonstrated consistent benefits across cognitive (EMACC, ISRL, GAS), neuropsychiatric (NPI), and biomarker (pTau217, GFAP) measures. Effect sizes ≥0.2 are considered clinically meaningful in early-phase trials. The convergent pattern across multiple mechanistically relevant endpoints supports biological activity of selective TNF inhibition in inflammation-enriched AD populations.

### Safety and Tolerability

XPro1595 was generally safe and well tolerated over 24 weeks, with no new safety signals and no deaths reported. The overall incidence of treatment-emergent adverse events (TEAEs) was similar between XPro1595 and placebo treated arms, with most events mild or moderate in severity (Table 3). The most common adverse event was injection site reactions (ISRs), occurring in ∼80% of XPro1595-treated participants versus 10% on placebo; ISRs were typically mild (redness or soreness), transient, and resolved without intervention. They occurred most frequently in the first 8 weeks and were in most instances regarded by participants and investigators as more of a nuisance than a safety concern. Twenty participants with ISRs underwent a 4-week dose modification where they received ¼ of the 1 mg/kg dose the first week with increasing ¼ doses until the participant reached the original study dose. Ten participants discontinued treatment due to ISRs, with a median discontinuation time of 35 days. Other adverse events occurring in ≥10% of either group included upper respiratory tract infections (placebo 16.4% vs 6.5% XPro1595) headache (placebo 10.4% vs 10.1% XPro1595), and arthralgia (placebo 6% vs 11.5% XPro1595) with no meaningful between-group differences. Serious adverse events were infrequent (placebo 7.5% vs 5.8% XPro1595), varied in type (e.g., transient ischemic attack, appendicitis, myocardial infarction in the XPro1595 group; syncope in placebo), and none were attributed to study drug. There were no severe or opportunistic infections, indicating that selective soluble TNF inhibition with XPro1595 did not cause immunosuppression. Laboratory parameters, including hematology and liver enzymes, showed no significant changes attributable to treatment, and there were no signs of hepatotoxicity or bone marrow suppression. Vital signs and ECGs remained stable, with no group differences in blood pressure or heart rate. Importantly, no participants required emergency evaluations or unscheduled neuroimaging for neurological events such as stroke, seizures, or visual disturbances, underscoring the favorable safety profile of XPro1595 in this trial.

**Table 3.** Summary of Adverse Events. Treatment-emergent adverse events (TEAEs) in the Safety Analysis Set (n=206). XPro1595 was generally well tolerated with no deaths reported. Injection site reactions were the most common adverse event in the active treatment group (79.9% vs 6.0% placebo), typically mild and transient. No opportunistic infections or immunosuppression signals were observed. Serious adverse events were infrequent and unrelated to study drug. The safety profile supports the selective mechanism of XPro1595 compared to broad TNF inhibitors.

| Event | Placebo<br>(n=67) | XPro1595<br>(n=139) | Total<br>(n=206) |
| --- | --- | --- | --- |
| Any TEAE | 59 (88.1%) | 131 (94.2%) | 190 (92.2%) |
| Any TEAE by Maximum Severity |  |  |  |
| Mild | 34 (50.7%) | 73 (52.5%) | 107 (51.9%) |
| Moderate | 22 (32.8%) | 56 (40.3%) | 78 (37.9%) |
| Severe | 3 (4.5%) | 2 (1.4%) | 5 (2.4%) |
| Any Serious TEAE | 5 (7.5%) | 8 (5.8%) | 13 (6.3%) |
| Any Treatment-Related Serious TEAE | 0 | 2 (1.4%) | 2 (1.0%) |
| Any TEAE of Special Interest (Hypersensitivity / Injection Site Reaction) | 7 (10.4%) | 112 (80.6%) | 119 (57.8%) |
| Any TEAE of Hypersensitivity | 3 (4.5%) | 43 (30.9%) | 46 (22.3%) |
| Any TEAE of Injection Site Reaction | 4 (6.0%) | 111 (79.9%) | 115 (55.8%) |
| Any TEAE Leading to Treatment Discontinuation | 2 (3.0%) | 12 (8.6%) | 14 (6.8%) |
| Any TEAE Leading to Study Withdrawal | 2 (3.0%) | 12 (8.6%) | 14 (6.8%) |
| Any TEAE with Fatal Outcome | 0 | 0 | 0 |
| **Adverse Events > 10% by System Organ Class & Preferred Term** |  |  |  |
| General disorders and administration site conditions |  |  |  |
| Any TEAE of Injection Site Reaction | 4 (6.0%) | 111 (79.9%) | 115 (55.8%) |
| Injection site reaction | 2 (3.0%) | 73 (52.5%) | 75 (36.4%) |
| Injection site erythema | 0 | 49 (35.3%) | 49 (23.8%) |
| Fatigue | 9 (13.4%) | 17 (12.2%) | 26 (12.6%) |
| Injection site hypersensitivity | 0 | 14 (10.1%) | 14 (6.8%) |
| Injection site pruritus | 0 | 12 (8.6%) | 12 (5.8%) |
| Infections and infestations |  |  |  |
| Upper respiratory tract infection | 11 (16.4%) | 9 (6.5%) | 20 (9.7%) |
| Musculoskeletal and connective tissue disorders |  |  |  |
| Arthralgia | 4 (6.0%) | 16 (11.5%) | 20 (9.7%) |
| Nervous system disorders |  |  |  |
| Headache | 7 (10.4%) | 14 (10.1%) | 21 (10.2%) |

No amyloid-related imaging abnormalities (ARIA-E or ARIA-H) were observed in any participant treated with XPro1595 (Table 4). Despite a high proportion of APOE ε4 carriers (∼70%, including 13% homozygous) and the inclusion of participants on anticoagulants, no cases of vasogenic edema or microhemorrhage occurred. Moreover, 34% of the SAF population entered the study with ≤14 cerebral microbleeds at baseline. The absence of ARIA is consistent with XPro1595’s distinct mechanism, targeting neuroinflammation, and represents a notable safety advantage, particularly in populations at elevated ARIA risk or in potential combination with amyloid-lowering therapies.

**Table 4.**
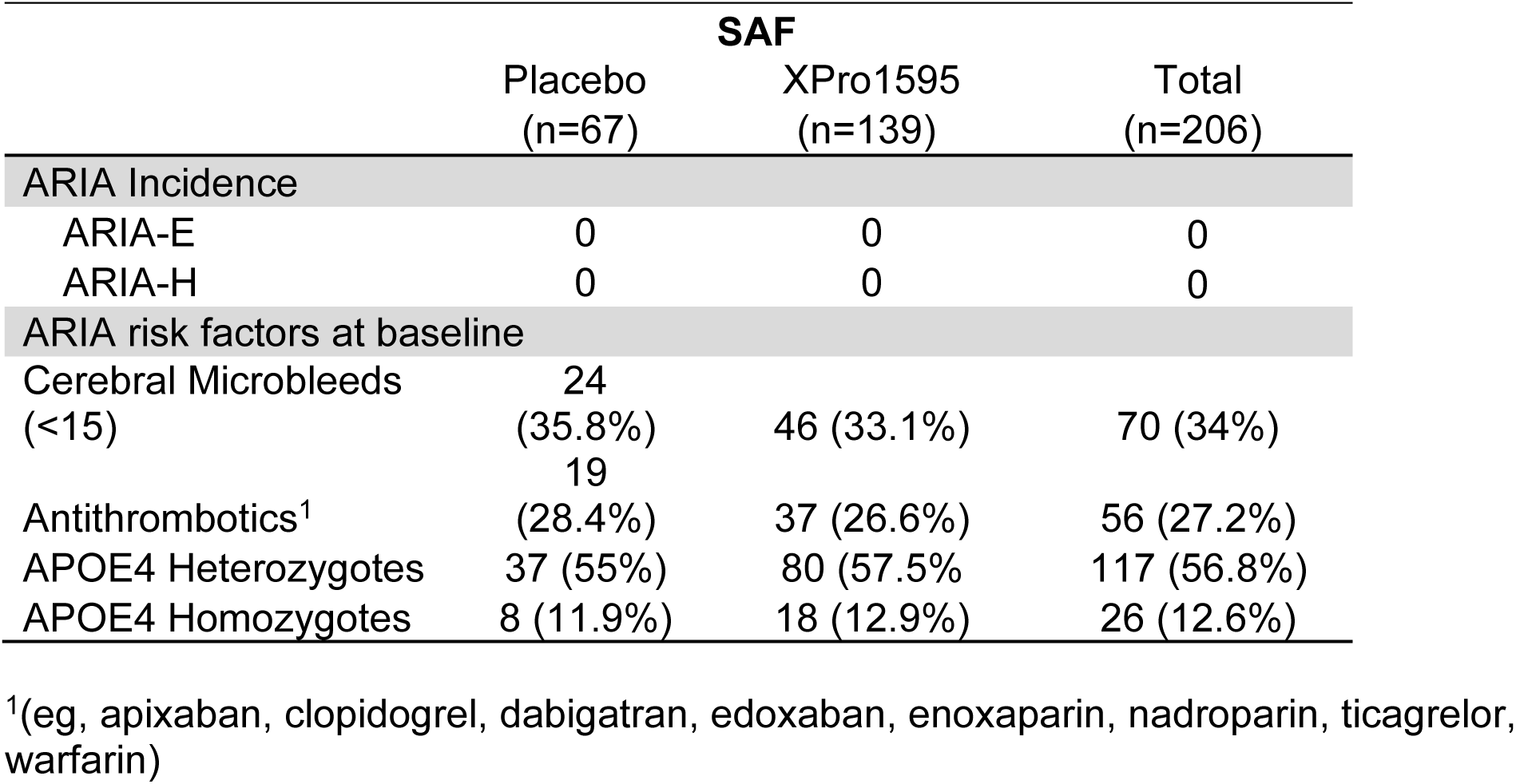
Amyloid-Related Imaging Abnormalities (ARIA) Assessment. ARIA incidence and baseline risk factors in the Safety Analysis Set. Notably, zero cases of ARIA-E (vasogenic edema) or ARIA-H (hemorrhage) were observed with XPro1595 treatment, despite enrollment of a high-risk population including ∼70% APOE ε4 carriers, 34% with baseline microbleeds, and 27% using antithrombotics. This finding contrasts markedly with approved anti-amyloid therapies and represents a significant safety advantage for XPro1595’s neuroinflammation-targeting approach.

In summary, the primary hypothesis that XPro1595 would show statistical superiority to placebo on the EMACC was not confirmed. Predefined population of amyloid positive patients with a high inflammatory burden, defined by at least two biomarkers of inflammation at baseline (ADi), revealed a consistent positive trend for XPro1595 on multiple assessments (EMACC, NPI, GAS, pTau217, and GFAP). A positive trend for treatment was also observed on the CDR-SB, NPI, pTau217 and GFAP in participants who received at least 21 mg/kg of the intended 23 mg/kg dose throughout the study (Fig. 3-5 and Table 2). Besides the frequently occurring injection site reactions, XPro1595 has a very favorable safety profile.

## Discussion

MINDFuL provides critical insights into selective soluble TNF inhibition as a potential therapeutic approach for Early Alzheimer’s disease. While XPro1595 did not show statistical significance on the primary endpoint within the overall study population, a prespecified subset of participants, enriched for the presence of amyloid and inflammatory biomarkers (ADi), demonstrated consistent trends indicating cognitive and neuropsychiatric benefits of XPro1595. This biomarker-enriched analysis, supported by comparable trends in changes in pTau217 and GFAP, reveals important mechanistic and clinical findings that warrant careful consideration for future development strategies.

### Primary Findings and Clinical Context

The absence of treatment effect in the mITT population was primarily driven by an unexpectedly stable placebo group, which contrasts with the anticipated cognitive decline typically observed in Early Alzheimer’s disease cohorts. This stability likely reflects the comparatively low disease burden in the MINDFuL cohort relative to participants in the anti-amyloid trials in Early Alzheimer’s disease (EAD). Specifically, baseline CDR-SB score in MINDFuL was 2.67 (1.3) whereas this value ranged from 3.2 to 3.3 (SD’s 1.34-1.5) in these trials, positioning MINDFuL participants at the milder end of the EAD disease spectrum^24^. Furthermore, 50 participants in the mITT group did not have confirmation of amyloid positivity. The inclusion of patients who turned out to have non-AD dementia likely impacted the mITT results by increasing variability and potentially confounding overall outcomes. Non-AD dementias, such as vascular or Lewy body dementia, are characterized by distinct underlying mechanisms, progression rates, and clinical features, which can further complicate interpretation of trial results and dilute the detection of treatment effects specific to AD^25^. EMACC, the primary endpoint, was empirically derived from cohorts of biomarker confirmed AD participants and has not been demonstrated to be fit for purpose to capture change in participants with non-AD dementia.

Within the biomarker-enriched population of amyloid-positive participants with elevated inflammatory markers (ADi, n=100), there were consistent trends favoring XPro1595 treatment such that participants treated with XPro1595 showed preserved cognitive skills, while ADi participants on placebo showed decline at 24 weeks. The estimated effect sizes for EMACC (Cohen’s d = 0.27 in ADi) indicated small-to-moderate advantages of XPro1595. This magnitude of benefit after only 6 months of treatment compares favorably to the magnitude of benefit observed on the ADAS-Cog at 18 months for approved anti-amyloid therapies in this population (of which the estimated effect sizes are between 0.10 and 0.15 for lecanemab in the CLARITY-AD trial^26^, and approximately 0.19 in the combined donanemab TRAILBLAZER ALZ-2 data^27^). The observed effect is especially notable as, by chance, placebo-treated participants had stronger cognitive skills at baseline than XPro1595-treated participants (ADi placebo EMACC mean score of 0.168 (SD 0.7) versus XPro1595 baseline mean of -0.073 (SD 0.7). The estimated effect sizes on EMACC were more robust when the Letter Fluency Test (LFT) component was excluded. Exploratory analyses of EMACC without the LFT were conducted due to translational concerns at multiple non-English–speaking sites and the lack of non-English speaking participants in the development sample. Although the precise impact of the LFT on the overall EMACC outcome remains unclear, future studies using EMACC in non-English–speaking populations should consider excluding the LFT to avoid this potential source of variability.

XPro1595 treated participants in the ADi population outperformed placebo treated participants on ISRL at week 24, demonstrating additional support for the cognitive benefit of XPro1595. ISRL, a delayed recall word list learning task, measures both acquisition and retention of new information, or episodic memory, a key cognitive deficit in AD.

These findings align with the hypothesis that participants with higher inflammatory burden may derive greater benefit from selective soluble TNF inhibition in shorter trials, as they demonstrate faster decline and thus provide better opportunities to detect treatment effects within a relatively short study window.

### Neuropsychiatric Benefits and Subfactor-Specific Effects

Secondary analyses revealed encouraging signals on neuropsychiatric symptoms, with XPro1595 demonstrating clinically relevant impact on agitation/hyperactivity (Cohen’s d = -0.37) in the ADi population. Neuropsychiatric symptoms often emerge early in the Alzheimer’s continuum and may be particularly sensitive to anti-inflammatory interventions in the short term, particularly in populations with preserved baseline function. The selective development of agitation/hyperactivity in placebo-, but not XPro1595-treated, participants suggests that XPro1595’s mechanism may have specific benefits for the development of neuropsychiatric symptoms in AD. This finding carries important clinical relevance given the established association between agitation in Alzheimer’s disease and accelerated cognitive decline, increased caregiver burden, and reduced quality of life for both those living with Alzheimer’s and their families.

### Functional Outcomes

Functional outcomes were assessed with the ECog and the functional domains of the CDR (Community Affairs, Home and Hobbies, and Personal Care). Neither instrument demonstrated differences between placebo and XPro1595-treated participants in the mITT population or in the ADi population. Interpretation of these findings requires consideration of the inherent limitations of informant-reported functional measures and the relatively short duration of the trial.

Functional assessments are indirect and rely on informants to detect and report changes in specific domains. Over a 24-week period, measurable differences are unlikely to emerge. For example, the ECog anchors judgments to changes relative to a decade earlier, limiting sensitivity to short-term fluctuations. Similarly, CDR functional ratings depend on informant reports and employ broad categorical scales (none, questionable/slight, mild, moderate, severe), which may obscure subtle changes. Individualized approaches such as the Goal Attainment Scale (GAS) may offer greater sensitivity to clinically meaningful functional changes; however, participants in this study were not required to select functional goals.

The EMACC was selected as the primary endpoint for MINDFuL because cognitive decline is an early manifestation of Alzheimer’s disease and reliably predicts subsequent functional deterioration^28–31^. The mild disease stage of the current sample, in conjunction with cognitive reserve, likely contributed to the minimal functional decline observed^30^. Consistent with this interpretation, effect sizes on CDR cognitive domains trended in favor of XPro1595, whereas CDR functional domains trended in the opposite direction.

Activities of daily living were also assessed with the ADCS-MCI-ADL and ADCS-ADL. However, substantial challenges limited interpretability. Few participants had consistent baseline and week 24 assessments across protocol versions, and ceiling effects were pronounced. At baseline, ≥80% of participants achieved maximum scores on at least half of the ADL items, restricting the capacity of these measures to detect longitudinal change.

### Mechanistic Insights and Target Engagement

Biomarker analyses strongly supported target engagement and biological activity. In the ADi population, XPro1595 attenuated the typical rise in both pTau217 and GFAP observed in placebo-treated participants, while showing a modest increase in plasma pTau217 in the broader mITT population. This divergent pattern suggests that there may be a minimum inflammatory burden necessary for an anti-inflammatory therapy (i.e. XPro1595) to effectively attenuate AD-related biomarkers. Indeed, a high inflammatory burden could indicate that inflammation may be the primary mechanism driving the person’s AD pathology. Finally, the parallel changes in neuroinflammation (GFAP) and neurodegeneration (pTau217) biomarkers provide mechanistic plausibility for observed clinical signals and demonstrate that selective soluble TNF inhibition can interrupt inflammatory cascades contributing to Alzheimer’s pathophysiology.

The differential treatment effects between study populations (mITT and ADi) provide compelling mechanistic validation of XPro1595’s selective approach to TNF inhibition. Unlike traditional TNF blockers that inhibit both soluble and transmembrane forms, XPro1595 specifically targets soluble TNF while preserving transmembrane TNF functions^8^ essential for immune homeostasis and glial cell protection. This mechanistic specificity appears most relevant in participants with a high inflammatory burden, as demonstrated by the ADi population’s distinct response profile.

### ARIA-Free Safety Profile: A Distinguished Advantage

The MINDFuL trial reported a complete absence of amyloid-related imaging abnormalities (ARIA) among participants treated with XPro1595, despite the deliberate enrollment of a high-risk population. Notably, >90% of ARIA events associated with anti-amyloid monoclonal antibodies occur within the first 24 weeks of exposure (Sperling et al., 2020), precisely the duration of this study, yet none were detected across participants that included ∼70% APOE ε4 carriers (13% homozygous), one-third with >14 baseline cerebral microbleeds, and 27% receiving antithrombotic therapy. The aggregation of such factors, each independently associated with elevated ARIA incidence, would be expected to amplify risk in a multiplicative fashion^32^. By contrast, ARIA represents clinically significant adverse event across all FDA-approved anti-amyloid monoclonal antibodies. For lecanemab, ARIA-E incidence is stratified by APOE ε4 status at 5.4% in non-carriers, 10.9% in heterozygotes, and 32.6% in homozygotes^26^. Donanemab demonstrates higher rates at 15.7%, 22.8%, and 40.6% across the same categories^33^, while aducanumab reports ARIA-E incidence ranging from 29.5% to 43.0% in APOE ε4 carriers depending on dose^34^. This disparity likely reflects mechanistic differences: amyloid antibodies activate microglia to clear plaques but can trigger maladaptive inflammation in primed immune systems, promoting vascular dysfunction and ARIA. In contrast, XPro1595 selectively neutralizes soluble TNF while sparing transmembrane TNF, rebalancing immune responses without overstimulation. This distinction avoids the ARIA-like complications seen with other immunostimulatory therapies such as TREM2 agonists^35,36^, underscoring the importance of restoring immune balance over broad activation.

XPro1595’s ARIA-free profile offers a unique advantage for combination strategies with amyloid-targeting agents. As combination approaches become essential in Alzheimer’s therapy, safety concerns from ARIA may limit the options that anti-amyloid treatments could form combinations with. XPro1595 could expand these possibilities by conditioning the brain through pre-treatment to normalize immune tone, by concurrent use to provide neuroprotection and suppress ARIA-inducing inflammation during amyloid clearance, or by sequential therapy to mitigate residual neuroinflammation afterward. Its dosing flexibility further supports these strategies by avoiding the tolerability constraints common with current anti-amyloid drugs.

### Disease Burden and Outcome Measure Sensitivity

The MINDFuL trial participants had lower disease baseline burden than participants in Early Alzheimer’s disease studies (e.g. ^26,27^). Specifically, more than 80% of participants had a CDR global score of 0.5, reflecting only “slight” concerns and “questionable” impairment. Episodic memory deficits were mild—an early and sensitive marker of Alzheimer’s pathology^22^, and 37% of the mITT sample would not have qualified for other trials that required impairments exceeding 1 standard deviation below age-adjusted norms. This relative preservation of function contributed to marked ceiling effects on functional measures, which was one potential limitation of the trial. On the MCI version of the ADL scale, which is tailored to less impaired populations, ≥80% of patients scored at ceiling on 10 of the 18 items. Such traditional instruments, developed to track decline across the entire Alzheimer’s spectrum, are not calibrated to detect change in very early-stage populations with substantial cognitive reserve and functional independence, especially within short trial periods.

The power analysis for the MINDFuL study was based on ADNI data, for which certain data was not available and may have influenced study outcomes. For example, change data was not available for letter fluency (a component of EMACC). Similarly, two of the four biomarkers - ESR, and HbA1C - were not available in ADNI and inflammatory decline was estimated by CRP and APOE4. While this approach represents a pragmatic application of robust natural history data commonly used in clinical trial design, these assumptions do not fully account for the complexity and potential variability inherent in disease progression trajectories.

The absence of detectable treatment effects may be attributed to fundamental differences between the modeling population and the actual study cohort. The ADNI participants used for power calculations demonstrated greater baseline cognitive and functional impairment compared to MINDFuL participants, who presented with milder disease severity and preserved functional independence. This population mismatch created a significant limitation in the ability to detect decline over the relatively brief 24-week observation period, despite implementing biomarker-based enrichment strategies targeting inflammatory markers that were hypothesized to accelerate disease progression.

Even modest deviations from the projected rate of decline could substantially impact study outcomes, particularly given the constrained 24-week observation window. The study design did not include formal power analyses for secondary functional endpoints, nor were sample size adjustments applied to these measures. Consequently, the failure to demonstrate measurable clinical decline among very early-stage participants—even those enriched for peripheral inflammatory biomarkers—represents an expected outcome rather than a study limitation. This finding underscores the inherent challenges in detecting meaningful clinical changes in populations with minimal baseline impairment over short-term follow-up periods.

### Dose-Response Relationships and Exposure-Efficacy Evidence

The MINDFuL trial trended important dose-exposure relationships supporting the therapeutic potential of XPro1595. Increased effect sizes with increasing drug exposure were observed across multiple endpoints in Dose Compliant Set analyses, which included participants receiving at least a total of 21 mg/kg (of 23 mg/kg) of study drug throughout the treatment period. The CDR-SB showed Cohen’s d values favoring XPro1595 increasing from -0.06 to -0.10 in the mITT population and from -0.05 to -0.13 in the ADi population with greater drug exposure. A similar result was observed in neuropsychiatric outcomes, with an effect size of 0.23 in ADi to 0.27 in participants within the ADi population who received at least 21 mg/kg throughout the study. These findings align with established clinical pharmacology principles and regulatory precedents, including FDA’s consideration of exposure-efficacy relationships in previous Alzheimer’s drug approvals, where clear dose-response patterns provided supportive evidence in overall efficacy results^37^.

### Future Development Strategy and Trial Design Considerations

The totality of the evidence revealed by the MINDFuL trial suggests a biologically and potentially clinically meaningful signal was detected even in the underpowered ADi cohort. These results support a focused development strategy leveraging both the ARIA-free safety profile and biomarker-driven efficacy signals. Future trials should prioritize enrollment of biomarker-enriched (amyloid positive with high inflammatory burden) populations while maximizing drug exposure to inform outcomes that appeared to be sensitive to cumulative drug exposure.

The contrasting results between analysis populations in MINDFuL highlight the critical importance of precision medicine approaches in Alzheimer’s therapeutic development, particularly in short trials. The ADi population closely represents the originally intended study population for which the trial was powered. The expansion of inclusion criteria during enrollment, due to operational constraints, likely diluted treatment signals in the broader analysis with participants lacking amyloid positivity and sufficient inflammatory burden, accounting for a lack of decline in the placebo group. Importantly, the operational challenges that forced protocol changes have been resolved, and the blood amyloid measurement infrastructure capacity has significantly improved globally since those early recruiting days of MINDFuL.

Several key considerations emerge for future neuroinflammation-targeted trials. The inflammatory biomarker enrichment strategy proved both feasible and biologically relevant, using readily available and clinically validated markers (hsCRP, ESR, HbA1C, APOE ε4) to identify populations more responsive to anti-inflammatory intervention, particularly within in a short time frame. The study demonstrated that the number of enrichment biomarkers at baseline was associated with treatment response, suggesting that overall inflammatory burden, rather than individual biomarkers or their combinations, determines the likelihood of response.

The EMACC composite, designed specifically for Early Alzheimer’s populations, demonstrated sensitivity to treatment effects in the enriched population that was not captured by traditional global measures. The composite’s empirical derivation and focus on cognitive domains most relevant to early disease may offer advantages over consensus-based instruments. However, the 24-week treatment period may have been insufficient for conventional functional measures to demonstrate meaningful change, particularly in early-stage populations with low clinical disease burden.

### Clinical Implications and Regulatory Pathway

The MINDFuL study results highlight XPro1595’s versatility as a therapeutic candidate with multiple potential applications: a standalone disease-modifying treatment for patients with biomarkers of inflammation, an add-on therapy for managing behavioral symptoms in AD, and/or a complementary treatment alongside amyloid-targeting therapies to mitigate inflammation and risk for ARIA. Continued development of XPro1595 as a disease-modifying treatment will benefit from identifying AD patients with elevated inflammatory markers and optimizing dosing to achieve meaningful functional improvements. Meanwhile, pursuing its development for the potential managing psychiatric symptoms addresses a critical gap in current AD care where treatment options remain severely limited.

The absence of ARIA represents a crucial differentiator in today’s treatment landscape, where safety concerns around cerebrovascular events restrict access for vulnerable populations, especially APOE ε4 carriers, who make up a significant proportion of AD patients and face elevated risks with current therapies^38,39^.

The regulatory pathway for XPro1595 may benefit from the precedent established by recent Alzheimer’s drug approvals, where biomarker evidence and dose-response relationships were considered supportive of efficacy claims. The mechanistic validation through biomarker changes, combined with the compelling safety profile and domain-specific clinical benefits, provides a strong foundation for continued development of XPro1595 in carefully selected populations with elevated inflammatory burden^37^.

## Conclusion

Based on the totality of the evidence observed, the MINDFuL study establishes XPro1595 as a promising therapeutic candidate that addresses critical gaps in Alzheimer’s treatment through its unique mechanism of immune modulation. Its ARIA-free safety profile enables treatment of high-risk patients typically excluded from amyloid-targeting therapies and opens new possibilities for combination strategies. While future development will require careful patient selection based on inflammatory biomarkers and optimized dosing regimens, the convergent evidence from cognitive, neuropsychiatric, and biomarker outcomes supports continued advancement of XPro1595 for both disease modification and symptomatic management in Alzheimer’s disease. These findings underscore the importance of precision medicine approaches in neuroinflammation-targeted therapies and highlight the potential for immune modulation to complement existing treatment paradigms.

## Author contributions

CJB, JJ, MB, MGT, SC, and RJT contributed to study design. CJB, JJ, MB, PP, KAS, SB, and LK analyzed and interpreted data. KAS and CJB wrote the manuscript. All authors contributed to writing and editing and approved the final manuscript.

## Acknowledgements

INmune Bio would like to acknowledge the study participants, their study partners, the investigators and all site staff, the CRO, all the vendors, and our colleagues who contributed to MINDFuL and made this trial possible. We are incredibly grateful for all their time and effort. This study received no funding.

