## Supplemental Data for "XPro1595, a Selective Soluble TNF Neutralizer, in Early Alzheimer’s Disease with Inflammation (ADi): Results from the Phase 2 MINDFuL Trial"

### Supplementary Data

### Supplementary Figure 1

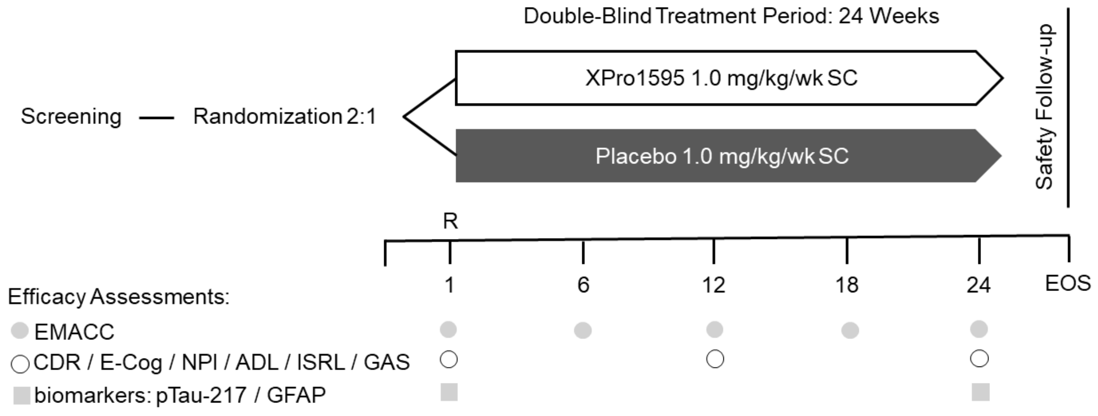


**Supplemental Fig. 1. Schematic overview of assessments during MINDFuL.** Participants were assessed by EMACC at randomization and every 6 weeks thereafter. Assessments for CDR, ECog, NPI, ADLs, ISRL, and GAS occurred at randomization and every 12 weeks thereafter. Biomarkers, pTau-217 and GFAP, were assessed at randomization and at 24 weeks. R = randomization. EOS = end of study.

### Supplementary Figure 2

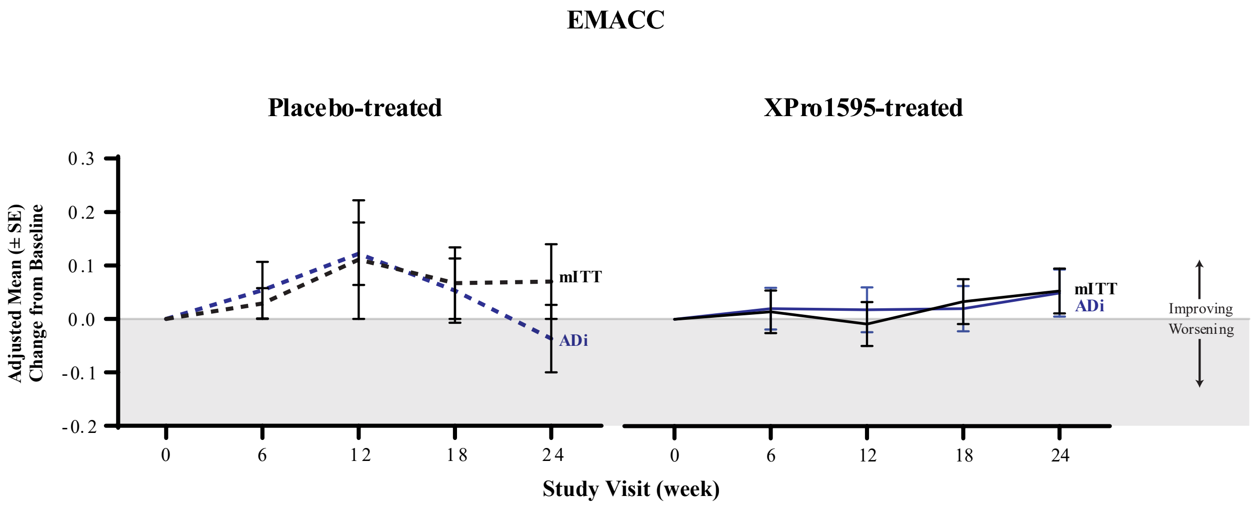


**Number of Participants**

| | | | | | | | | | | |
|---|---|---|---|---|---|---|---|---|---|---|
| **mITT** | 66 | 64 | 64 | 56 | 58 | 134 | 123 | 115 | 109 | 111 |
| **ADi** | 28 | 27 | 26 | 23 | 23 | 72 | 66 | 62 | 60 | 61 |

**Supplemental Fig. 2. EMACC Performance by Placebo and XPro1595 treatment in mITT and ADi.** Detailed trajectory analysis showing cognitive performance over 24 weeks in placebo- and XPro1595-treated participants in mITT and ADi. Placebo treated participants in the ADi group exhibited the anticipated decline characteristic of Early AD progression, while placebo-treated participants in mITT showed relative stability whereas XPro1595-treated participants showed similar change over time regardless of study population. Data are presented as LSM +/- SEM. Negative scores reflect disease worsening.

Supplementary Table 1

**Supplemental Table 1. Complete Inclusion and Exclusion Criteria.** Comprehensive listing of eligibility criteria for the final protocol version. Key inclusion criteria included Early AD diagnosis (MCI or mild dementia), CDR global score 0.5-1.0, MMSE >22, and presence of inflammatory biomarkers (hsCRP >1.5 mg/L, ESR >10 mm/h, HbA1C >6%, or APOE ε4 carrier status). Major exclusions included other causes of cognitive impairment, recent immunosuppressive therapy, and contraindications to MRI. The criteria were designed to enroll participants most likely to benefit from anti-inflammatory intervention.

| **Inclusion criteria** | **Exclusion criteria** |
|---|---|
| To be eligible for study entry, patients must satisfy all of the following criteria:<br>1. Adult patients 50 years to ≤ 85 years of age at the time of consent;<br>2. Meets the diagnostic criteria of MCI of probable Alzheimer's disease (Jack et al. 2018; NIA- AA) or mild dementia as clinically described in McKhann, (2011) and corresponding to stages 3 or 4 of | Patients will be excluded from the study if 1 or more of the following criteria are applicable:<br>1. Have any contraindications to MRI scanning, including cardiac pacemaker/defibrillator, ferromagnetic metal implants (e.g., in-skull and cardiac devices other than those approved as safe for use in MRI scanners).<br>2. Have any evidence of other clinically significant lesion(s) that |

| | |
|---|---|
| the revised AD staging system (Jack, 2018). (NIA - AA);<br>3. Amyloid positive (documented in medical history or assessed during screening through blood test);<br>4. CDR global rating at screening of 0.5 or 1;<br>5. MMSE > 22;<br>6. ECog memory subscale items mean > 1.5;<br>7. Presence of at least 1 inflammatory biomarker:<br>o · hsCRP > 1.5 mg/L<br>o · ESR > 10 mm/h<br>o · HbA1C > 6DCCT%<br>o · At least 1 APOE4 allele (documented in medical history or assessed during screening through blood test)<br>8. Consents to APOE genotyping; | could confound or indicate a dementia diagnosis other than AD on brain CT and/or MRI at Screening.<br>· Exhibit other significant pathological findings at the Screening MRI including but not limited to: an area of cortical superficial siderosis; evidence of cerebral vasogenic edema (ARIA-E); macro-hemorrhage(s) (>10mm); multiple (>15) microhemorrhages (ARIA-H); one large (>15 mm on axial plane) infarct or multiple (≥2) lacunes (as defined by STRIVE consensus criteria); an area of encephalomalacia suspect of prior head trauma; aneurysm(s); subdural hematoma; severe small vessel or diffuse white matter disease (defined as Fazekas scale score of 3); any space occupying lesion(s), or brain tumor(s), also including infective lesions (however, lesions diagnosed as meningiomas or arachnoid cysts and less than 1 cm at their greatest |

| | |
|---|---|
| 9. Modified Hachinski Ischemic Score ≤ 4 (see Section 9.8 for additional details);<br>10. Either currently or previously (in pre-AD condition) literate and capable of reading, writing, and communicating effectively with others;<br>11. Must have a sixth-grade education or work experience to exclude developmental causes of cognitive dysfunction;<br>12. Residence in an assisted living is allowed as is personal assistances provided in the home, however at time of enrollment participant must be able to perform most ADL with minimal assistance, and participant must be permitted sufficient independence to allow assessment of change in ADL;<br>13. May be using the following concomitant medications for | diameter need not be exclusionary). Radiological evidence for another dementia etiology (such as stroke, traumatic brain injury, Lewy Body disease, substance/medication use, CNS infection, Prion disease, Parkinson’s disease, Huntington’s disease, etc.) major brain diseases, or other CNS trauma, that interfere with the participant's ability to comply, and dementia-related disease as determined by clinical diagnosis.<br>3. Receives considerable help to carry out basic ADL living either in the home or as a resident in a nursing home or similar facility.<br>4. Lifetime history of a major psychiatric disorder including schizophrenia and bipolar disorder. Major depressive disorder that has resulted in 2 or more hospitalizations in a lifetime. Major depressive episode during the past 5 years that is judged by the |

| | |
|---|---|
| management of AD at Screening and during the study, including cholinesterase inhibitors (donepezil, rivastigmine, and galantamine), glutamate inhibitors (memantine), and multi-modal treatments (Namzaric®). Other, non-immunosuppressive therapies such as medications to treat behavior symptoms including: suvorexant (insomnia), SSRIs for depression and anxiety, and drugs for narcolepsy (Xyrem® or modafinil) may also be used and should be discussed with the sponsor. These medications must be started at least 90 days before the first dose of XPro1595 and the regimens must remain constant throughout the study.<br>14. The patient must be willing and able to provide informed consent | clinical team unlikely to have been part of Alzheimer's prodrome. History of suicidal behavior, or answer of 'yes" to C-SSRS suicidal ideation items 4 or 5 within 12 months of screening.<br>5. History of substance abuse within 12 months; use of cannabis or cannabis products within 24 weeks of consent.<br>6. Have taken within the last 90 days from Day 1: corticosteroids or other immunosuppressive drugs, thalidomide or other TNF active drugs, minocycline, first- or second-generation antipsychotics (e.g., aripripozole) or aducanumab or other anti-amyloid therapies. Topical corticosteroids for cutaneous, nasal, or ocular applications can be used with approval from Medical Monitor. Patients taking cholinesterase |

| | |
|---|---|
| prior to any study procedures being performed.<br>15. Has a study partner for the duration of the trial who either lives in the same household or interacts with the patient at least 4 hours per day and on at least 4 days per week, who is knowledgeable about the patient's daytime and night-time behaviors and who can be available to attend all clinic visits in person at which study partner assessments are performed. This study partner should agree to monitor and report on concomitant medications, understand the study requirements, and assist the participant in meeting study requirements. Patients with study partners that interact with the patient on at least 4 days per week but for less than 4 hours per day who the investigator determines as | inhibitors, memantine, or antidepressant medication for less than 90 days from Day 1 (i.e., must be on stable dose for at least 90 days prior to Day 1).<br>7. Approval by the Medical Monitor is required if a patient takes any drug known to alter cognitive functioning. Examples include benzodiazepines, sedating antihistamines, anticonvulsants, sedating antidepressants (e.g., tricyclic antidepressants), anticholinergics (benztropine), antiparkinsonian medications (amantadine, selegiline, benztropine), psychostimulants, narcotic medications within 4 weeks of baseline.<br>8. Any untreated infection or any infection that the PI deems is not currently controlled with current therapies. |

| | |
|---|---|
| able to provide an adequate assessment of the patient may also participate with prior approval from the Sponsor.<br>16. All male subjects who are sexually active with a female of childbearing potential (FCBP) must agree to use a highly effective method of contraception during the treatment period and until 90 days after the last dose of treatment. All females of childbearing potential (FCBP) must have a negative urine pregnancy test and agree to use two highly effective method of contraception (including at least one barrier method) during the treatment period and 30 days after the last dose of treatment. | 9. Enrolled in another clinical trial where patients receive treatment with an investigational drug or treatment device or have had previous treatment with any investigational medicinal product within 60 days or 5 half-lives (whichever is longer) prior to study drug treatment.<br>10. A prior organ or stem cell transplant.<br>11. A major adverse cardiac event within 24 weeks before consent.<br>12. Lymphoma, leukemia, or any malignancy within the past 5 years with the exception of malignancies with negligible risk of metastasis or death, such as basal cell or squamous cell carcinomas of the skin or cervical carcinoma in situ that have been resected with no evidence of metastatic disease for 3 years. |

| | |
|---|---|
| | 13. Jaundice, active hepatitis, or known hepatobiliary disease (except asymptomatic cholelithiasis).<br>14. Positive screening assessment for viral HBsAg or HCV antibody and positive HCV RNA or HIV, or a history of illicit drug injecting.<br>15. Seated blood pressure of ≥ 165/105 mmHg at Screening.<br>16. Unable to comply with the study procedures and assessments.<br>17. Known hypersensitivity to investigational product or its excipients.<br>18. Any investigator site personnel directly affiliated with this study and their immediate families. Immediate family is defined as a spouse, parent, child, or sibling, whether biological or legally adopted. |

| | |
|---|---|
| | 19. Those who are committed to an institution by virtue of an order issued either by the judicial or the administrative authorities. |

### Supplementary Table 2

**Supplemental Table 2. Protocol Evolution and Enrollment by Version.** Timeline of protocol amendments and their impact on participant enrollment. Initial versions required 2+ inflammatory biomarkers and mandatory amyloid confirmation, but operational constraints led to modification of these criteria (Version 5-6). The final versions restored amyloid requirements while maintaining single biomarker enrichment. This evolution explains the mixed population in mITT analyses and underscores the importance of biomarker-driven enrollment for inflammation-targeted therapies.

| MINDFuL's protocol version | Key inclusion / exclusion criteria or changes | Number of enrolled participants |
|---|---|---|
| Version 1 | Inclusion: mAD, 2 or more biomarkers of inflammation, amyloid-positive, 60-85 years old | 0 |
| Version 2 | Inclusion: mAD, 2 or more biomarkers of inflammation, amyloid-positive, 60-85 years old | 7 |
| Version 3 | Inclusion: mAD, 1 or more biomarkers of inflammation, amyloid-positive, 60-85 years old | 0 |
| Version 4 | Inclusion: mAD, 1 or more biomarkers of inflammation, amyloid-positive, 60-85 years old | 5 |

| | | |
|---|---|---|
| Version 5 | Inclusion: mAD, 1 or more biomarkers of inflammation, no amyloid-positivity required, 60-85 years old | 51 |
| Version 6 | Inclusion: mAD or MCI, 1 or more biomarker of inflammation, no amyloid-positivity required, 50-85 years old, ECog memory domain >1.5 | 53 |
| Version 7 | Inclusion: mAD or MCI, 1 or more biomarker of inflammation, amyloid-positivity required, 50-85 years old, ECog memory domain >1.5 | 88 |
| Version 8 | Inclusion: mAD or MCI, 1 or more biomarker of inflammation, amyloid-positive, 50-85 years old, ECog memory domain >1.5 | 2 |
